# Evaluating Digital Device Technology in Alzheimer’s Disease via Artificial Intelligence

**DOI:** 10.1101/2021.11.07.21265705

**Authors:** Meemansa Sood, Mohamed Aborageh, Daniel Domingo-Fernández, Robbert Harms, Thomas Lordick, Colin Birkenbihl, Andrew P Owens, Neva Coello, Vaibhav A. Narayan, Dag Aarsland, Maximilian Bügler, Holger Fröhlich, for the Alzheimer’s Disease Neuroimaging Initiative, RADAR-AD Consortium

## Abstract

The use of digital technologies may help to diagnose Alzheimer’s Disease (AD) at the pre-symptomatic stage. However, before implementation into clinical practice, digital measures (DMs) need to be evaluated for their diagnostic benefit compared to established questionnaire-based assessments, such as the Mini-Mental State Examination (MMSE) for cognition and Functional Activity Questionnaire (FAQ) for daily functioning. Moreover, the quantitative and qualitative relationship of DMs to these well understood scores needs to be clarified to aid interpretation. In this work we analyzed data from 148 subjects, 58 cognitively normal and 90 at different stages of the disease, which had performed a smartphone based virtual reality game to assess cognitive function. In addition, we used clinical data from Alzheimer’s Disease Neuroimaging Initiative (ADNI). We employed an Artificial Intelligence (AI) based approach to elucidate the relationship of DMs to questionnaire-based cognition and functional activity scores. In addition, we used Machine Learning (ML) and statistical methods to assess the diagnostic benefit of DMs compared to questionnaire-based scores. We found non-trivial relationships between DMs, MMSE, and FAQ which can be visualized as a complex network. DMs, in particular those reflecting scores of individual tasks in the virtual reality game, showed a better ability to discriminate between different stages of the disease than questionnaire-based methods. Our results indicate that DMs have the potential to act as a crucial measure in the early diagnosis and staging of AD.

## Introduction

Alzheimer’s disease (AD) is an irreversible chronic neurodegenerative disorder^1^ and the leading cause of dementia^2,3^ representing 60-70% of the cases^4^. With the lack of definitive cure for the disease, it has become essential to target the disease at an early stage^5^. A prerequisite for the differential diagnosis of disease stages of AD is a distinctive pattern of cognitive and functional impairment^6,7,8^. Cognitive impairment is examined through questionnaire-based tests that assess multiple cognitive domains, including attention, memory, language, concentration etc, e.g., the Mini-Mental State Examination (MMSE)^9^. Functional impairment is measured by tests like the Functional Activity Questionnaire (FAQ), which is marked by deteriorations in activities of daily living (ADLs), such as use of technology, managing finances, and shopping^10,11,12^. However, questionnaire-based assessments only provide a subjective snapshot of a patient’s cognitive and functional abilities, which can vary over time. Hence, digital device technologies, including smartphone apps, currently receive increasing interest in the assessment of dementia symptomatology^13,14^.

These technologies can measure features of disease symptoms remotely and deliver real-time data to healthcare providers^15^. Digital measures (DMs), for example, scores reached in an augmented reality (AR) game, could allow for an accurate, quantitative, and objective monitoring of disease symptoms^16^. However, before any clinical use, DMs have to be evaluated by assessing their relationship to established clinical scores and understanding their diagnostic benefit. To this end, the (Innovative Medicines Initiative) IMI project RADAR-AD (www.radar-ad.org) follows the ambition to evaluate a broad panel of digital technologies with respect to their potential for early AD diagnosis while focusing on ADLs.

As an example of a panel of digital technology, in this work we focused on a digital medical device built by Altoida, Inc.^17^ that deploys a battery of immersive AR and motor activities over iOS, and Android smartphones and tablets, including activities which require the user to place and find objects in a virtual environment. The activities are designed to put the user under a cognitive load representative of what they experience when performing complex ADLs. The device generates several scores, including performance in individual tasks and derived overall neurocognitive domain scores (backed by DSM-5 criteria) that can be used to identify cognitive impairment and neurocognitive disease. The overall ambition of the work presented in this paper was two-fold:

1. Understanding the relationship between the DMs produced by the Altoida app and established tests and questionnaires (e.g., MMSE, FAQ).
2. Assessing the ability of DMs to discriminate between cognitively normal (CN) and mild cognitively impaired (MCI) subjects in comparison to an established cognitive screening test.

## Results

Tables 1, and 2 provides an overview about both employed cohorts (Altoida, ADNI) in terms of baseline summary statistics grouped by diagnostic stages. Patients in ADNI are on average longer educated and older than Altoida patients. In ADNI a high fraction of patients within the MCI and demented groups are carriers of at least one APOE4 risk allele (MCI: 52%, dementia: 69%).

**TABLE 1.**
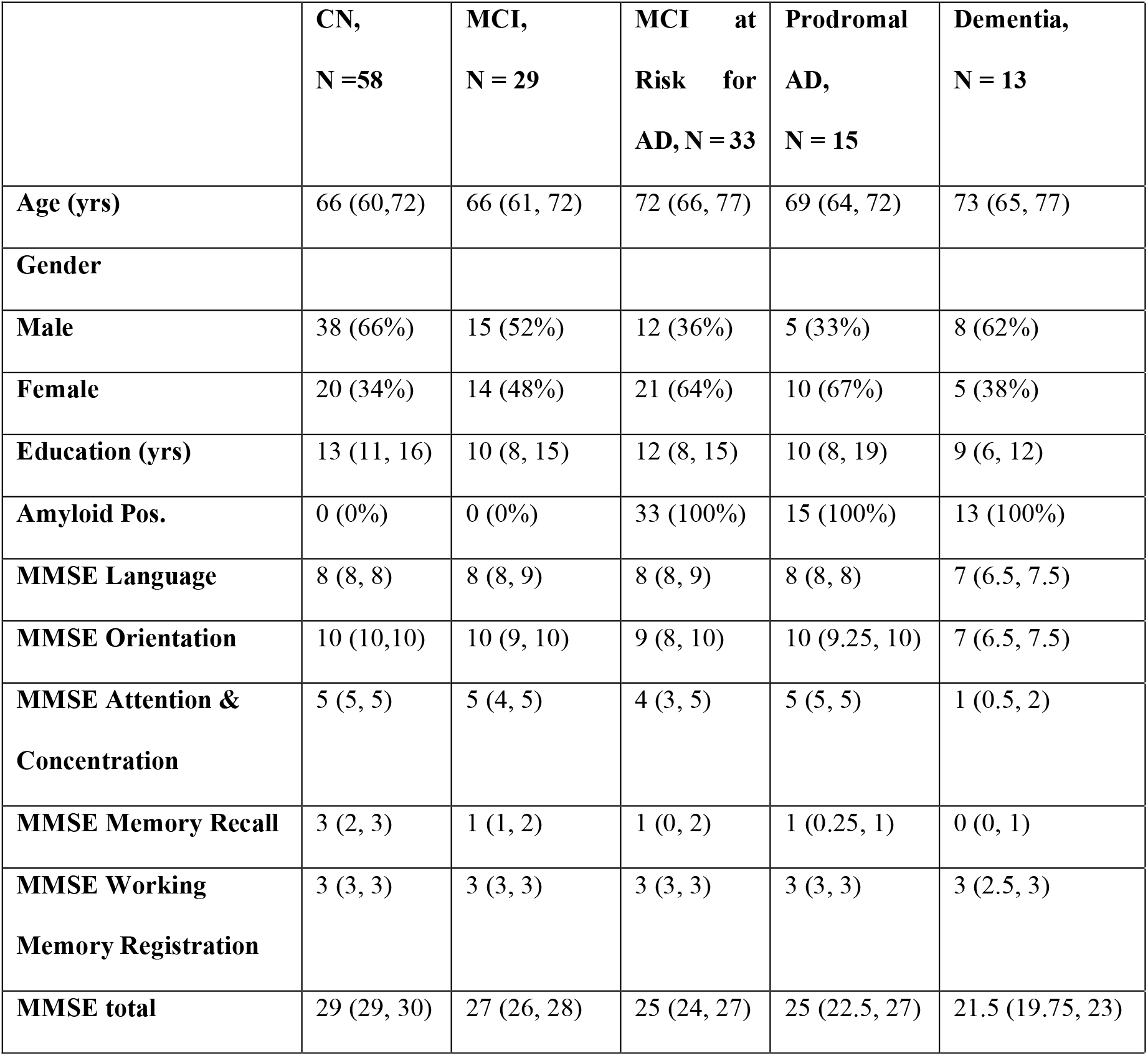
Summary statistics of Altoida demographic and amyloid data grouped by diagnostic stages, data are n (%), or median (IQR in brackets), unless specified otherwise.

**TABLE 2.**
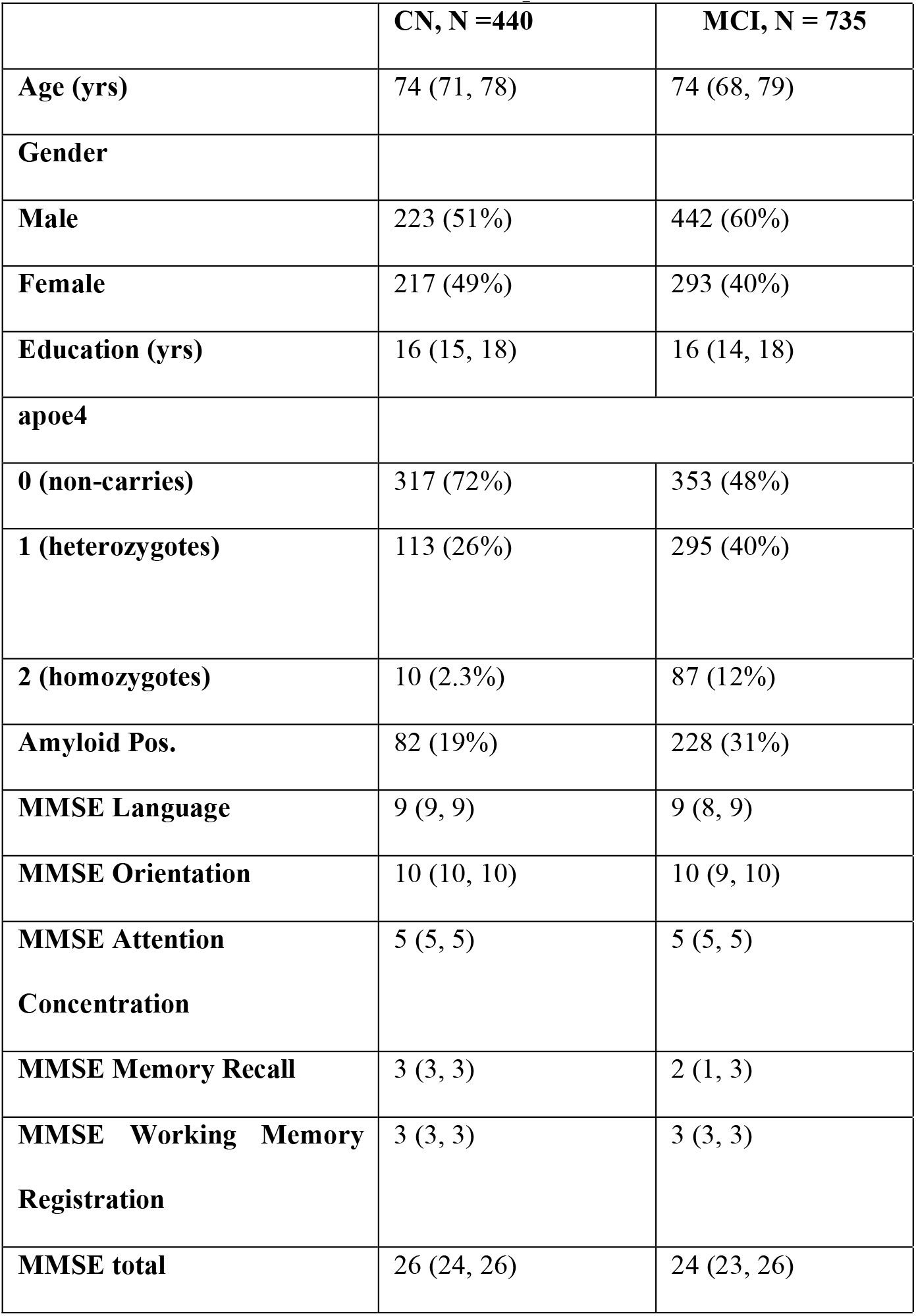
Summary statistics of ADNI demographic data, amyloid and APOE4 status grouped by diagnostic stages, data are n (%), or median (IQR in brackets), unless specified otherwise.

|  | **CN, N =440** | **MCI, N = 735** |
| --- | --- | --- |
| **Age (yrs)** | 74 (71, 78) | 74 (68, 79) |
| **Gender** |  |  |
| **Male** | 223 (51%) | 442 (60%) |
| **Female** | 217 (49%) | 293 (40%) |
| **Education (yrs)** | 16 (15, 18) | 16 (14, 18) |
| **apoe4** |  |  |
| **0 (non-carries)** | 317 (72%) | 353 (48%) |
| **1 (heterozygotes)** | 113 (26%) | 295 (40%) |
| **2 (homozygotes)** | 10 (2.3%) | 87 (12%) |
| **Amyloid Pos.** | 82 (19%) | 228 (31%) |
| **MMSE Language** | 9 (9, 9) | 9 (8, 9) |
| **MMSE Orientation** | 10 (10, 10) | 10 (9, 10) |
| **MMSE Attention**<br>**Concentration** | 5 (5, 5) | 5 (5, 5) |
| **MMSE Memory Recall** | 3 (3, 3) | 2 (1, 3) |
| **MMSE Working Memory**<br>**Registration** | 3 (3, 3) | 3 (3, 3) |
| **MMSE total** | 26 (24, 26) | 24 (23, 26) |

### Overview about Data from Altoida Inc

Altoida’s digital medical device was used to collect data from 148 subjects. Patients were diagnosed into different stages of the disease according to guidelines of the revised National Institute on Aging (NIA) and Alzheimer’s Association (AA)^18^, see supplements section 2.7. The smartphone application combines data from hand micro-movements™ and microerrors™, gait micro-errors™, visuospatial navigation micro-errors™, and recent voice parameters. One complete session consists of various motor function activities, a series of complex AR activities, and speech analysis^17^ (details in Supplements Section S2).

Sensor data generated from these tests is considered as “raw” and using feature extraction, we can generate DMs. These measures were subsequently used to assess the overall cognitive performance of a subject via several cognitive domain scores related to perceptual motor coordination, complex attention, cognitive processing speed, inhibition, flexibility, visual perception, planning, prospective memory and spatial memory (Section S2.2). For our analysis, we examined 148 subjects, out of which 123 subjects were measured once, 20 subjects were measured twice, and 5 subjects were measured thrice, leading to a total of 178 subject records. In addition to DMs, for all 148 subjects traditional MMSE scores were available, which are often used in clinical routine.

### Overview about Data from Alzheimer’s Disease Neuroimaging Initiative (ADNI (https://adni.loni.usc.edu/))

We examined 1445 subjects, having longitudinal measurements at baseline, months 6, 18, 24 and 36. The data consisted of two different diagnostic stages:

i. CN
ii. MCI. MCI has been defined in ADNI as follows^19^:1) subjective memory complaints reported by study participant, study partner, or clinician; 2) memory loss according to an education-adjusted WMS-R Logical Memory Test; 3) global Clinical Dementia Rating (CDR) score of 0.5; and 4) general cognitive and functional performance sufficiently preserved such that a diagnosis of dementia could not be made.

We considered MMSE and FAQ subitem scores as cognitive tests (Tables S1, S2), because i) they are also assessed in the Altoida data (MMSE), ii) reflect ADL (FAQ) and iii) generally have been suggested to reflect disease progression^20, 21, 22^.

In addition to MMSE, we used neuroimaging derived features, cerebrospinal fluid (CSF) protein measurements (amyloid-beta, tau, and phospho-tau), and genetic burden on a collection of AD specific disease mechanisms / gene sets retrieved from the NeuroMMSig database^23^ (Supplements Section S1, Table S4).

We determined the amyloid status of the subjects based on the neuroimaging features, Florbetapir (AV45) amyloid PET value of the subjects, (described in Table S4). If subjects had an AV45 value >1.11, they were amyloid positive and negative otherwise^24^.

Patients in ADNI are on average longer educated and older than Altoida patients. In ADNI a high fraction of patients within the MCI and demented groups are carriers of at least one APOE4 risk allele (MCI: 52%, dementia: 69%).

### Non-trivial dependencies between DMs, MMSE scores and diagnostic state in Altoida data

Our analysis of Altoida data resulted into a quantitative network between different groups of variables, which is depicted in Figure 1. Numbers on edges indicate the level of statistical confidence, i.e., a higher value means a stronger support by the data for the existence of the respective connection.

**FIGURE 1.**
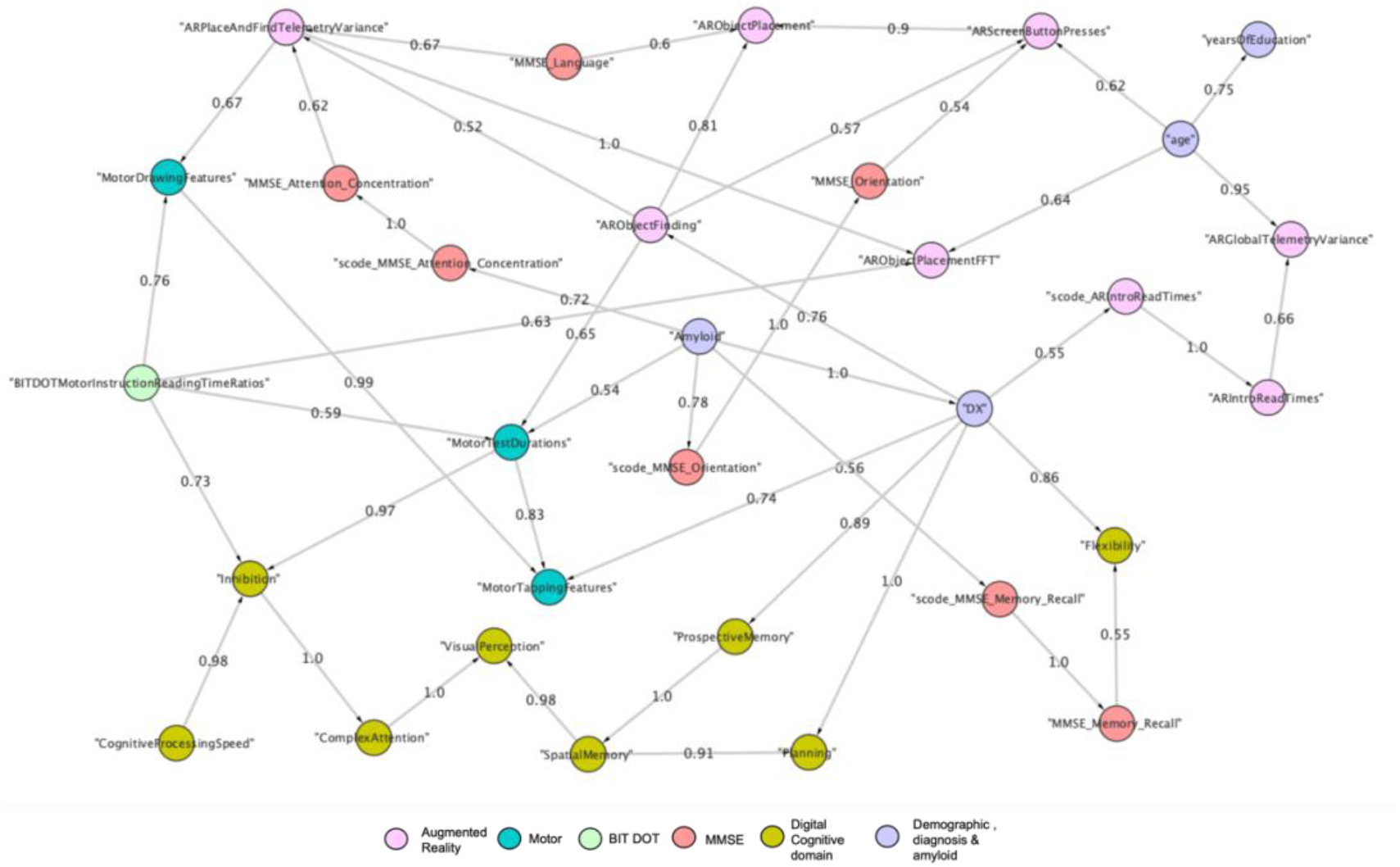
Non-trivial dependencies between DMs, MMSE scores and diagnostic state in Altoida data. \The depicted network represents dependencies between variables and variable groups learned from Altoida data (a part of the network is represented here). Numbers on edges indicate the level of statistical confidence (bootstrap probability >0.5). A higher value indicates a higher confidence in the existence of a connection. Nodes are color coded by the group they belong to. Nodes isolated from the rest of the network are not shown.

According to our model, the MMSE sub-domain, “Orientation” is linked to the pressure and accuracy of touch screen button pressing in the AR game (Spearman rank correlation *ρ* = −0.47, 95% CI [-0.67, −0.21], adj. p < 0.0001). Spearman rank correlation is a measure between −1 and 1, where −1 indicates a perfect anti-correlation and +1 a perfect positive correlation. MMSE language subdomain is connected to the telemetry variance observed in object placing and finding (*ρ* = - 0.60, 95% CI [-0.76, −0.38], adj. p < 0.0001) and to the score derived from tasks associated with placing the object in the virtual environment (*ρ* = 0.34, 95% CI [0.06, 0.58], adj. p < 0.05). We also observed that the MMSE sub-domain associated with memory recall is connected to the digital cognitive domain that measures the ability to switch between thinking about two different concepts (*ρ* = 0.31, 95% CI [0.02, 0.55], adj. p < 0.05). The corresponding scatter plots for all these connections are illustrated in Figure S2. Overall, our model revealed non-trivial dependencies between DMs and MMSE scores, which are far away from simple one-to-one relationships.

A point of further interest was the dependency of all DMs on the diagnosis, which was reflected via corresponding paths in our Variational Autoencoder Modular Bayesian Network (VAMBN) network. We verified these dependencies by conducting Mann-Whitney U-tests of individual DMs between MCI and CN patient groups, which demonstrated highly significant differences of the cognitive domains, “Cognitive Processing Speed” (speed and accuracy of information processing), “Prospective Memory” (ability to remember to carry out intended actions in the future) and “Spatial Memory” (ability to recognize items that previously appeared in physical space) between MCI and CN patients, see Table S8. In addition, individual digital task scores demonstrated significant differences between both patient groups (Table S9). Likewise, we performed the analysis for MMSE subitem scores, which demonstrated significance in all cases, except for the working memory registration task, which was unconnected to the diagnosis in our VAMBN network (Table S10). All DMs showed a significant age dependency (Table S11).

### Non-trivial dependencies of DMs, MMSE scores, FAQ scores and diagnostic state predicted in ADNI data

ADNI data analysis resulted in a rather large quantitative network model, which can be explored interactively via the web-based tool DigiAD that we developed for this purpose (https://digi-ad-viewer.scai.fraunhofer.de). An excerpt of the network is depicted in Figure 2. Importantly, DMs in this network had been predicted for each individual ADNI patient using the AI model trained on Altoida data. The prediction used the diagnostic state, age, education, gender and MMSE subitem scores of each patient, see Section 2.3.2. Accordingly, we confirmed most connections between MMSE subdomains and DMs, which we had previously also observed in the Altoida data, see Table 3 for details. The corresponding scatter plots are shown in Figure S3. Furthermore, Table S12 highlights the expected dependencies of DMs on age.

**FIGURE 2.**
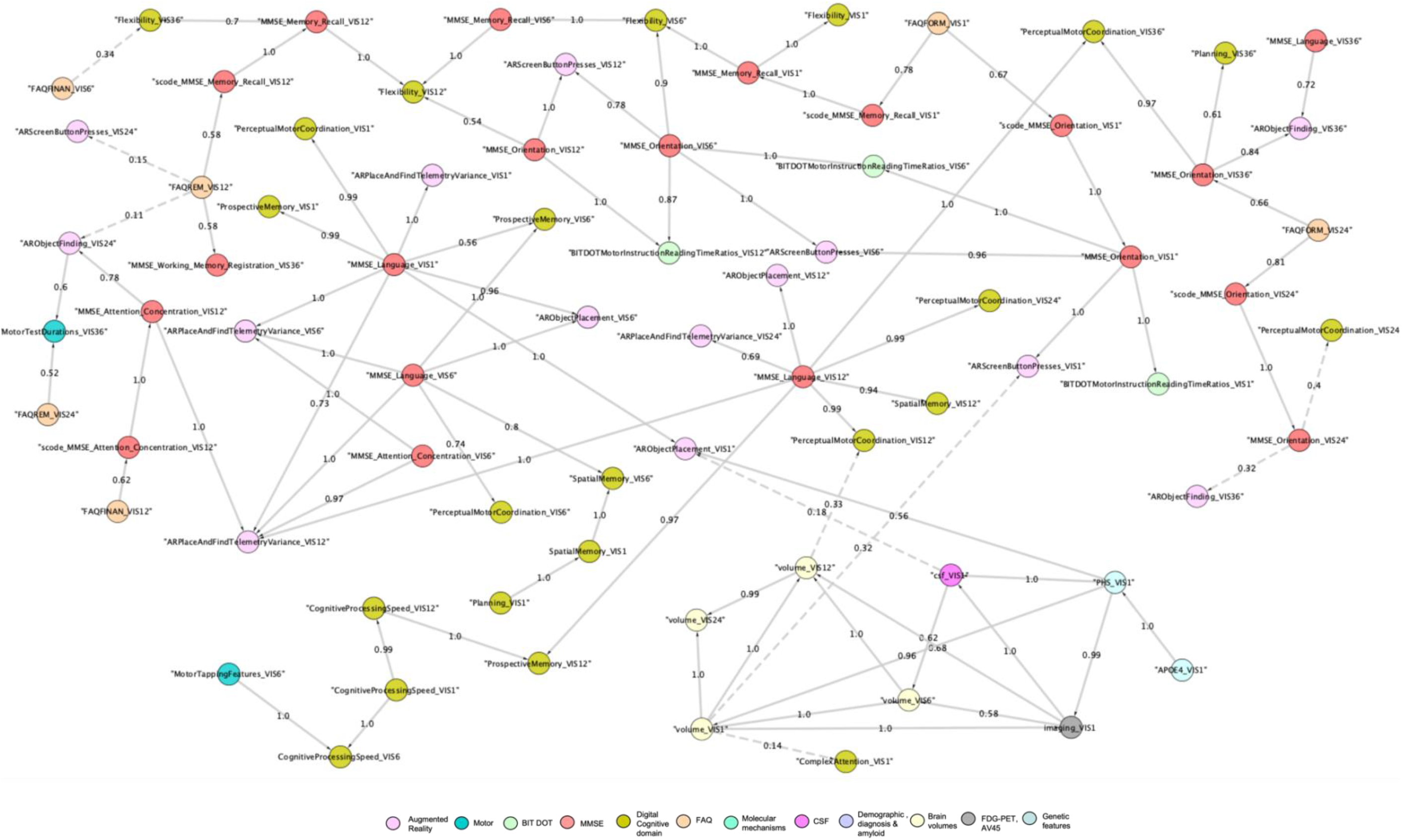
Non-trivial dependencies of DMs, MMSE scores, FAQ scores and diagnostic state predicted in ADNI data. The depicted network represents dependencies between variables and variable groups learned from ADNI data. Numbers on edges indicate the level of statistical confidence (bootstrap probability >0.5). A higher value indicates a higher confidence in the existence of a connection. Nodes are color coded by the group they belong to. Here shown a part of the model depicting interesting connections between clinical features and digital features. Solid edges represent a bootstrap probability > 0.5, dashed edges a bootstrap probability < 0.5. Nodes isolated from the rest of the network are not shown.

**TABLE 3.** Spearman Rank Correlation between MMSE subdomains and DMs in ADNI with confidence intervals (CI) and adjusted p values (Holm’s method)

| MMSE sub domain (from node) | Digital Measure (to node) | Spearman Rank correlation coefficient ($\rho$) | CI [lower ci, upper ci] | adj.p |
| --- | --- | --- | --- | --- |
| MMSE Orientation (baseline) | AR Screen Button Presses (baseline) | -0.36 | [-0.47, -0.25] | <0.0001 |
| MMSE Orientation (month 6) | AR Screen Button Presses (month 6) | -0.47 | [-0.56, -0.37] | <0.0001 |
| MMSE Orientation (month 12) | AR Screen Button Presses (month 12) | -0.41 | [-0.51, -0.3] | <0.0001 |
| MMSE Language (baseline) | AR Place and Find Telemetry Variance (baseline) | -0.60 | [-0.67, -0.51] | < 0.0001 |
| MMSE Language (month 6) | AR Place and Find Telemetry Variance (month 6) | -0.53 | [-0.62, -0.43] | < 0.0001 |
| MMSE Language (month 12) | AR Place and Find Telemetry Variance (month 12) | -0.62 | [-0.70, -0.54] | < 0.0001 |
| MMSE Language (baseline) | AR Object Placement (baseline) | 0.35 | [0.23,0.46] | <0.0001 |
| MMSE Language (month 6) | AR Object Placement (month 6) | 0.18 | [0.05,0.29] | <0.0001 |
| MMSE Language (month 12) | AR Object Placement (month 12) | 0.19 | [0.07,0.31] | <0.0001 |
| MMSE Memory Recall (baseline) | Flexibility (baseline) | 0.66 | [0.59,0.73] | <0.0001 |
| MMSE Memory Recall (month 6) | Flexibility (month 6) | 0.54 | [0.45,0.62] | <0.0001 |
| MMSE Memory Recall (month 12) | Flexibility (month 12) | 0.49 | [0.39,0.58] | <0.0001 |

In addition to these expected and confirmed findings, we also predicted new dependencies, for example between the “orientation” domain of MMSE and the difference in time when reading the instructions to the motor tests for the second time (*ρ* = −0.20, 95% CI [-0.32, −0.08], adj. p < 0.0001) at baseline. Additionally, dependencies were predicted between the “language” domain of MMSE and the motor coordination in response to perceived input (perceptual motor coordination) (*ρ* = - 0.24, 95% CI [-0.36, −0.12], adj. p < 0.0001).

We also found novel predicted dependencies between DMs and FAQ subitems. Note that FAQ was not assessed in the Altoida data. There was a direct link from the FAQ measure related to “remembering appointments, family occasions, holidays, medications” at month 24 to digital task of the total duration of each motor test at month 36 (*ρ* = 0.14, 95% CI [0.02, 0.26], adj p = <0.005). Another example was the indirect relationship between the same subdomain of FAQ at month 12 and the cognitive domain “Flexibility” (*ρ* = −0.24, 95% CI [-0.36, −0.12], adj. p <0.0001). Corresponding scatter plots along with other examples are illustrated in Figures S4 and S5. Interestingly, Our VAMBN model also suggested direct associations of DMs with CSF biomarkers, brain volumes and molecular mechanisms, but these connections were either not found statistically stable or not significant.

We repeated the same analysis using only those feature modalities from ADNI, which were also present in Altoida (i.e., demographic features, MMSE scores, DMs). This generally confirmed the same connections between DMs and MMSE subitem scores discussed before (Figure S6).

### Diagnostic benefit of DMs compared to questionnaire-based outcomes

To evaluate the potential diagnostic benefit of DMs compared to established clinical outcome measures we trained and evaluated machine learning classifiers to discriminate between patient records at CN (n = 62) versus MCI (n = 81) stage. Our results on Altoida demonstrated no significant increase in the prediction performance on held out test data when using cognitive domains in addition to MMSE subitem scores (p = 0.9373, Wilcoxon signed rank test) compared to only using MMSE scores (Figure 3A). However, when using individual digital tasks in addition to MMSE scores a strongly significant improvement was found (p<0.0001, Wilcoxon signed rank test). Similarly, when using digital tasks alone the prediction performance was significantly better compared to only using MMSE scores (p<0.0001, Wilcoxon signed rank test). Hence, digital task scores seem to offer a more accurate diagnosis compared to MMSE subitem scores. Figure 3B depicts a full Receiver Operator Characteristic (ROC) curve of the classifier using only digital task scores, demonstrating that these measures offer a ∼80% sensitivity at a 10% false positive rate. To better understand the relevance of individual digital tasks in comparison to MMSE subitem scores, we explored the absolute values of the coefficients in the CN vs. MCI classifier using DMs plus MMSE scores. The most relevant digital task scores were related to object placement and object finding (Figure 3C). Altogether, DMs contributed 80.3% of the overall feature importance, whereas MMSE subitem scores contributed less than 20% (Figure 3D).

**FIGURE 3.**
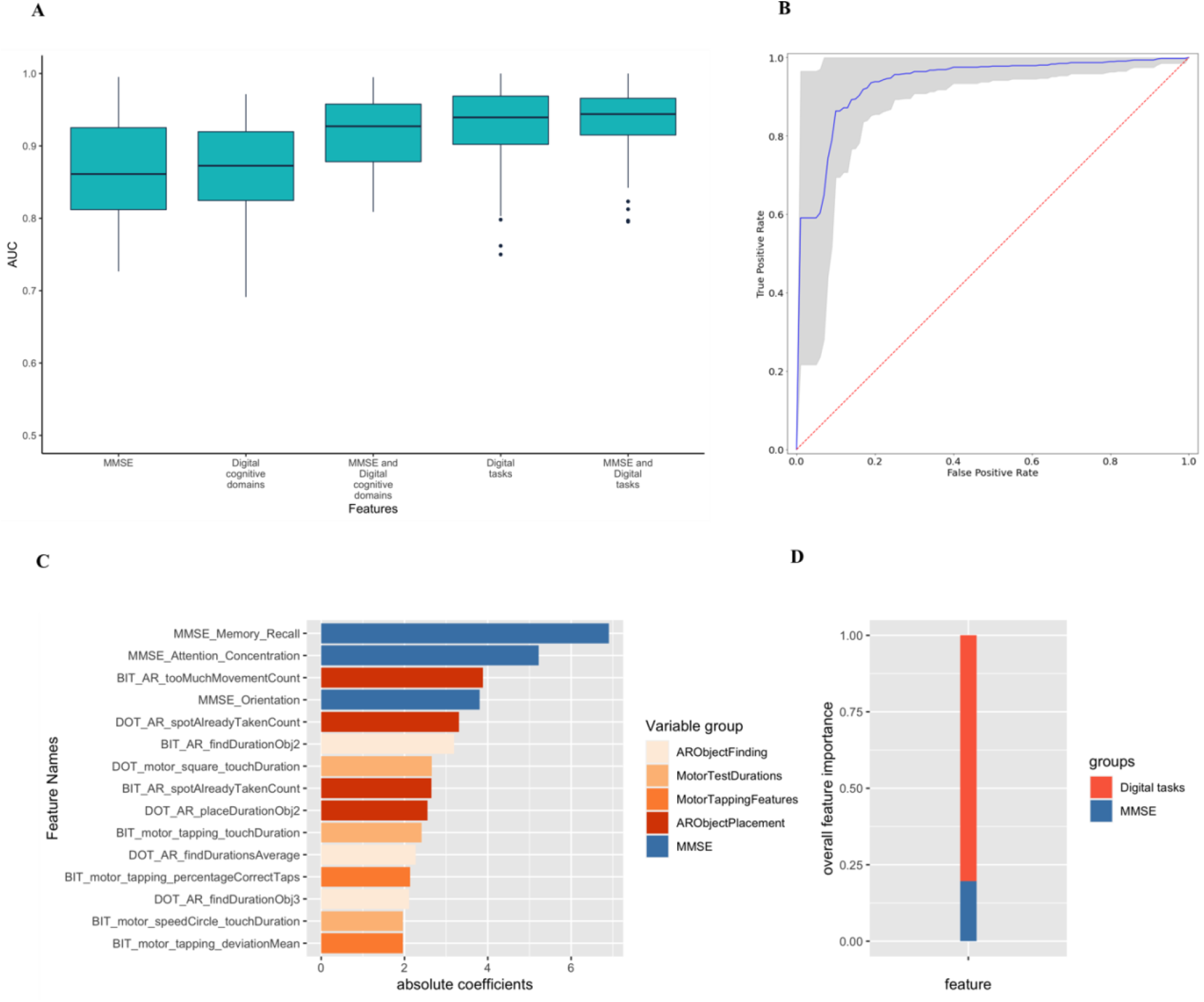
A: Performance of a sparse group lasso trained on different modalities of data to classify subjects into CN and MCI using Altoida data, measured via the area under ROC curve (AUC-ROC). The boxplots show the distribution of the AUC-ROC on held out test data, measured via 10 times repeated 5-fold repeated cross-validation. B: Performance of a sparse group lasso trained on only digital tasks, depicting mean and confidence interval of the AUC-ROC on held out test data, measured via 10 times repeated 5-fold repeated cross-validation. C: Feature importance when using MMSE plus individual digital tasks observed in Altoida data (measured by the absolute value of coefficients) using a machine learning classifier discriminating between CN and MCI patients. Only the top 15 features are shown. The complete list can be found in Supplement Table S13. D: Overall feature importance of MMSE versus digital tasks, measured by the sum of absolute coefficient values. The total sum of all absolute coefficient values is normalized to 1.

In comparison to MMSE, FAQ subitem scores better reflect ADL. Our analysis on ADNI data showed that FAQ scores better discriminate between CN and MCI than MMSE scores (Supplementary Section 10). However, the increase in prediction performance is only 2% on average, compared to 7% when using DMs instead of MMSE scores. This suggests a diagnostic benefit of DMs also in comparison to questionnaire-based ADL assessments. Notably, ADNI does not contain DMs (although we were able to predict them) and thus an equivalent evaluation to Altoida was not possible in a reliable manner.

As a last step we evaluated whether the DMs discriminating between CN and MCI patients (i.e., the predictors in the classifier only containing DMs) would also demonstrate a statistically significant difference between MCI and other disease stages (prodromal AD, dementia). Statistically ssignificant differences in cognitive domains, “Flexibility”, “Planning” “Prospective Memory” and “Spatial Memory” were observed between MCI and prodromal AD patients (Table S8). Similarly, between MCI and dementia the cognitive domains “Perceptual Motor Coordination”, “Complex Attention”, “Flexibility”, and “Planning” showed significant differences. In addition, several digital task scores demonstrated significant differences between these groups (Table S9).

We repeated the same analysis with MMSE subitem scores (i.e., the predictors in the classifier only containing MMSE scores) and we did not find a statistically significant difference in MMSE score between MCI and prodromal AD. Between MCI and dementia all MMSE subitem scores except for “memory recall” were significant (Table S10).

## Discussions

Digital biomarkers currently receive a lot of attention, because they have the potential for a more objective, robust, and sensitive measurement of disease symptoms compared to traditional questionnaire-based assessments. In addition, digital biomarkers can be used in outpatient situations and thus allow for a continuous monitoring of patients in their natural environment. In the AD field, the potential use of digital biomarkers for objective assessment of cognitive impairment has been discussed by several authors^17,25^, including the possibility for early disease diagnosis^26,27^. Specifically digital biomarkers reflecting activities of daily living have been suggested to enable an early disease diagnosis^16^. However, the actual diagnostic benefit compared to traditional questionnaire-based assessments needs to be critically evaluated in order to justify the use in clinical routine. In this context, it is important to remark that digital biomarkers comprise a set of abstract features (DMs), which represent properties of a signal measured via a digital device. The interpretation of those features is thus not always clear. Hence, there is a need to better understand (quantitatively) the relationship of DMs to clinical questionnaires.

Based on a dataset of 148 subjects, in this work we were able to disentangle the apparently non-trivial and complex relationships that exist between different types of DMs reflecting performance across distinct neurocognitive domains, as well as individual tasks obtained from a virtual reality game with questionnaire-based assessments of cognition (MMSE, FAQ). Our identified associations were statistically stable, significant, and predictable. Our analysis hence provided a very deep and novel level of interpretation of DMs. In addition to interpretation of DMs, our analysis showed that digital cognitive domains and digital task scores measured via the Altoida app offer a higher accuracy and sensitivity for predicting the disease at an early stage than questionnaire-based tests. Notably, DMs demonstrated significant differences between MCI and prodromal AD, whereas MMSE subitem scores were not significant. In this context, our analysis on the Altoida data revealed that digital task scores generally seem to be clearly better suited for a discrimination between CN and MCI patients than digital cognitive domain scores. They offer a higher diagnostic accuracy than questionnaire based MMSE scores. Relevant digital task scores were related to object placement and finding in the virtual environment, to motor tapping and to the duration of motor tests. According to our analysis on ADNI, DMs are also suggested to have a benefit compared to questionnaire-based FAQ assessments for predicting the disease stage, hence better reflecting ADL.

The main limitation of our work is the limited sample size of the Altoida dataset, the cross-sectional design and lack of neuropsychological tests other than MMSE. On the other hand, ADNI contains a rich collection of neuropsychological tests measured longitudinally, but no DMs. Correlations observed between DMs and classical outcome measures such as MMSE were statistically significant but with a moderate strength. It remains a subject of future research to validate our findings using further AD cohorts measuring both DMs and questionnaire-based assessments of cognition.

Altogether, the approach described in this paper could serve as a blueprint to better understand DMs in AD and to evaluate their diagnostic accuracy compared to traditional questionnaire-based scores. This is in turn a pre-requisite for moving DMs from an evaluation phase into actual use in clinical studies and clinical routine.

## Methods

### Overview about analysis strategy

The overall strategy of our approach is outlined in Figure 4.

**FIGURE 4.**
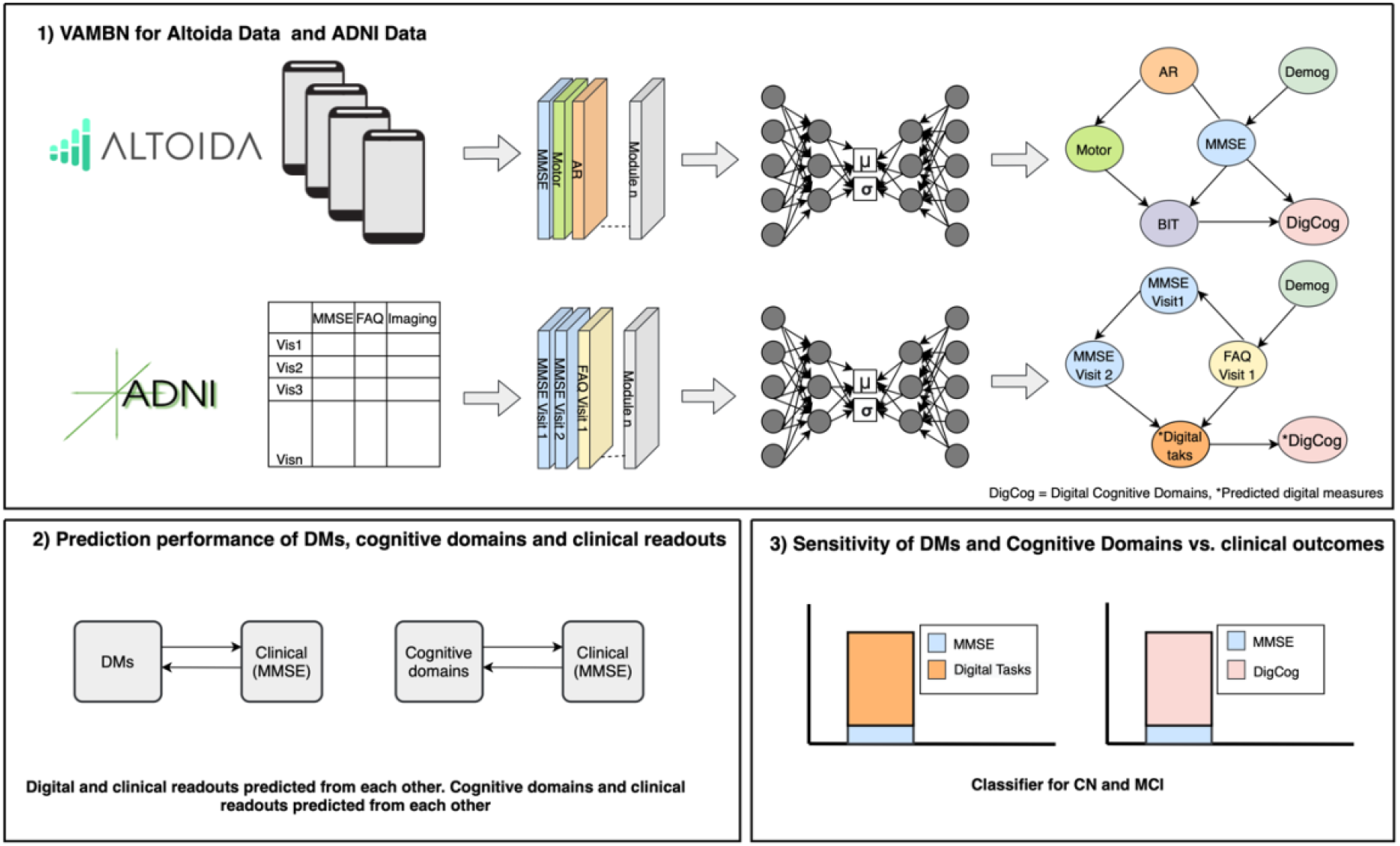
Brief workflow of our methodology is described here in 3 steps. 1) Fitting the VAMBN model of Altoida and ADNI. 2) Testing the prediction performance of DMs from clinical measures like MMSE (Mini Mental State Examination) and vice versa 3) Testing the classifier on Altoida for classifying disease into CN (cognitively normal) and MCI (Mild Cognitive Impairment).

### Fitting a VAMBN model

The Altoida app results into 11 individual scores for different tasks in the virtual environment (Table S3) and there are 9 derived scores for different cognitive domains (Section S2.2). Hence, there is a very large number of potential correlations between the Altoida DMs and the 5 different MMSE subitem scores (Table S1). To address this issue, in this paper we employed an AI based approach, which results into a quantitative and visualizable network structure. More specifically, we employed the VAMBN^28^ approach. Fitting a VAMBN model consists of several steps:

1. Definition of interpretable variable groups in the data. For example, DMs and motor scores were grouped into different modules. Demographic features (e.g., age, gender etc.) were not grouped into a module as we wanted to see their individual effects on other features. Modules and their description are presented in Tables S3 and S4.
2. Learning of a low-dimensional representation of each module. This was achieved by training a Heterogenous Incomplete Variational Autoencoder^29^. The result was a Gaussian Mixture Model encoding the higher dimensional input data.
3. Learning of an augmented Bayesian Network (BN) connecting the modules. The BN contained additional auxiliary variables to account for missing values in the data, e.g., due to patient drop-out. We repeated BN learning 1000 times using random subsamples of the data to derive a measure of statistical confidence^30^.

The result of steps 1. - 3. was a quantitative network model representing patient-level data. Details about the learning procedure and evaluation of the fit of the model are provided in the supplementary material section 4.

### Evaluating the predictability of digital endpoints

A ready-trained VAMBN model can be used to infer the value of a specific variable based on the value of other variables within an individual patient. In our case, we used this approach to assess the predictability of DMs from other nodes. More specifically, we randomly split the entire data into 10 folds and sequentially left out one-fold for testing while training the augmented BN on the rest of the data (10-fold cross-validation). The same procedure was repeated 10 times. Prediction performances were compared against those of a standard Random Forest regression model^31^, see details in supplements section 5.1. Estimated normalized root mean square error (NRMSE) were below 0.20 units for most of the features indicating a good prediction performance. Afterwards, we used the VAMBN model trained on the entire Altoida data to infer DMs in ADNI using the common features that were observed in both the datasets (diagnosis, demographics (age, education, gender), MMSE subitems). Technical details are provided in the supplements section 5.2.

### Assessment of the ability of digital measures to discriminate different diagnostic stages

The last step of our analysis focused on the question, whether DMs could better discriminate between CN and MCI than questionnaire-based assessments like MMSE. For this purpose, we trained a machine learning classifier (sparse group lasso)^32^ to differentiate between the CN vs MCI stage using the Altoida data.

In the classification task, the dependent variable indicated the patient subgroup (CN, MCI), whereas the independent / predictor variables were MMSE subitem scores digital tasks and digital cognitive domains. In Altoida data, a total of 143 patient records were used to train a classifier out of which 62 patient records were in normal state and 81 patient records were in either MCI or MCI at risk for AD.

The following combinations of scores from Altoida data were considered to build a classifier:

1. Neurocognitive domain scores derived from the Altoida’s app (termed as “digital cognitive domains”) plus MMSE subitem scores
2. Scores for individual tasks in the virtual reality game (termed as “digital tasks”) plus MMSE subitem scores
3. MMSE subitem scores alone
4. Digital cognitive domains alone
5. Digital tasks alone

Prediction performance of each machine learning model was evaluated by 10 times repeated 5-fold cross-validation. That means, the entire dataset was randomly split into 5 folds, and one of these folds was subsequently left out for testing the model, while the rest of the data was used for training. Details about the training and evaluation procedure can be found in the supplements. The result of the performance evaluation was the area under curve-receiving operating characteristics (AUC-ROC) as a measure of prediction performance. The AUC-ROC is a measure between 0 and 1, where 1 indicates a model that will classify the subjects perfectly into CN and MCI and 0.5 a model that classifies no better than chance^33^. To assess the variability of the AUC-ROC on the random splitting of the entire dataset into folds, the overall 5-fold cross-validation procedure was repeated 10 times.

After performance evaluation, the model was trained on the entire data and the coefficients for each predictor variable were extracted to assess their statistical influence. We were able to identify a set of discriminative DMs, and those DMs were subsequently tested for statistical significance in comparison of MCI patients vs. prodromal AD (n = 15) and fully demented patients (n = 13) using a Mann-Whitney U-test. Multiple testing correction was performed via Holm’s method. We also developed a classifier based on the same strategy for baseline ADNI subjects to compare the prediction performance of FAQ and MMSE to classify subjects into CN and MCI. Details are described in supplementary section 10.2 and 10.3.

## Supporting information

Supplementary_file_updated

## Data Availability

The respective data owners of the data used in this publication provided access to the datasets. For ADNI, approval was received by applying for data access on http://adni.loni.usc.edu/data-samples/access-data/ web server. For Altoida, approval to use the data was provided by it’s data owners.

## Acknowledgements

The research leading to these results has received partial support from the Innovative Medicines Initiative Joint Undertaking under grant agreement #IMI2-2017-12-01, resources of which are composed of financial contribution from the European Union’s Horizon 2020 Framework Program and EFPIA companies’ in-kind contribution.

The authors would like to thank Luise Gootjes-Dreesbach, Tamara Raschka, and Martin Hofmann-Apitius for helpful discussions and Helena Balabin and Sumit Madan for occasional technical support.

## Author Contributions

Initiated, designed, and supervised the project: HF, MB; guided the project: HF, MB, DA, AW, NC; compiled the data: RH; MS implemented the methods and ran the experiments. TL helped in the generation the genetic burden scores. MA, CB helped with the development and analysis of classifiers. DDF developed the DigiAD web application. MS and HF drafted the manuscript.

