## Supplementary_file_updated for "Evaluating Digital Device Technology in Alzheimer’s Disease via Artificial Intelligence"

† Alzheimer's Disease Neuroimaging Initiative: Data used in the preparation of this article were obtained from the Alzheimer's Disease Neuroimaging Initiative (ADNI) database (adni.loni.usc.edu). As such, the investigators within the ADNI contributed to the design and implementation of ADNI and/or provided data but did not participate in analysis or writing of this report. A complete listing of ADNI investigators can be found at http://adni.loni. usc.edu/wp-content/uploads/how_to_apply/ ADNI_Acknowledgement_List.pdf

### 1. Encoding of SNPs on Molecular Mechanism Level

The information related to SNP (single nucleotide polymorphism) data was downloaded from the ADNI server (http://adni.loni.usc.edu/data-samples/access-data/) for different subsets of data:
ADNI 1: 581,500 SNPs from 757 subjects, measured via Illumina Human610-Quad Bead Chip platform
ADNI 2/GO: 708,870 SNPs from 432 subjects, measured via Illumina HumanOmniExpress
ADNI 3: 16,743,712 SNPs from 327 subjects measured via Illumina Omni 2.5M.

SNPs were imputed via the Michigan Imputation Server[1] using the Haplotype Reference Consortium (HRC) reference panel, which consists of 64,976 haplotypes[2]. SNPs were considered as reliable imputed, if the $r^2$ was above 0.3 (default setting).

Two major databases, PheWAS Catalog[3] and DisGeNet[4] were used to gather AD associated SNPs. SNPs collected from both these databases were further extended by those SNPs, which were in strong in Linkage Disequilibrium ($r^2 > 0.8$) via HaploReg (Version 4.1)[5].

In order to map these SNPs to genes, two steps were performed:

- Mapping to genes in closest chromosomal location via HaploReg and using default settings.
- Via expression quantitative trait loci (eQTL) mapping using gene expression data from brain tissues obtained from the GTEx Portal (GTEx Consortium, 2013). Only cis-eQTL were taken into account.

Subsequently, the NeuroMMSig knowledge base[6] was used to find those genes that can be linked to AD-related biological mechanisms. As we required a minimal number of mechanisms, we selected 20 relevant/well-known mechanisms that also had a large number of genes in its corresponding network. Then, the corresponding SNPs were mapped to each of this mechanism.
The number of SNPs available in each of this mechanism are illustrated in Table S2. Each of these mechanisms (except one) was encoded as a module. That means, a HI-VAE encoder was trained to map to a lower dimensional representation. Details about HI-VAE training are explained in Section 4 of this document.

### 2. Information on Altoida Data: Concepts and Terminology

#### 2.1. The Altoida test

The Altoida test is a purely smartphone-sensor-based, digital biomarker-based prediction model, which only includes age, sex and years of education to personalize NMI on an individual level. Using a tablet or smartphone device, a person is asked to perform a series of tasks ranging from simple motoric tasks to complex Augmented Reality (AR) tasks. During these tasks the handheld device collects telemetry and touch data from the built-in sensors, enabling profiling of hand micromovements, screen touch pressures, walking speed, navigation trajectory, cognitive processing speed and more. The motor activities consist of drawing activities and tapping activities. In the shape drawing activity, the subject is asked to draw various shapes (on the touch screen) using their index finger. In the tapping activity, the subject is first asked to tap a simple series of buttons (left, right) and then a similar series in which buttons are randomly highlighted. During these motor activities, eye tracking can be enabled to get more sensor data. In the AR activities, the subject is asked to walk around the room holding the device in their hands in front of them. On the screen the environment is shown, augmented with digital objects. The subject is asked to virtually place three objects by clicking a "place" button on the screen while holding the device near to a physical surface such as a table or desk. Afterwards, the subject is asked to find these three objects by holding the tablet close to the location where they placed the objects. A speech activity can then be (optionally) performed in which the subject is asked to verbally describe an image. All activities above are performed twice in a specific order: motor, AR, speech, motor, AR, speech. For more details on the complete activity battery, please refer to Bugler et al. 2020.

#### 2.2. Cognitive domains

The cognitive performance of a subject ranks the subject's test performance relative to his/her peers. The performance is presented as a set of scores over several cognitive domains. To compute, the recorded sensor data from an Altoida test is reduced into a set of characteristic elements, or data features. For each of these features, a percentile score is computed by comparing the subject's feature value to those of a group of healthy subjects of the same age and gender. Per domain, a specific set of these percentile scores are combined to form each of the cognitive domain scores.

Altoida currently reports on the following cognitive domains:

- *Perceptual Motor Coordination*, motor coordination in response to perceived input
- *Complex Attention*, capacity to choose what to pay attention to and what to ignore
- *Cognitive Processing Speed*, speed and accuracy of information processing
- *Inhibition*, ability to tune out stimuli that are irrelevant to the task
- *Flexibility Ability*, to switch between thinking about two different concepts
- *Visual Perception*, visual search speed, visual perception and efficiency
- *Planning*, process of thinking about the activities required to achieve a desired goal
- *Prospective Memory*, ability to remember to carry out intended actions in the future
- *Spatial Memory*, ability to recognize items that previously appeared in physical space

Altoida is currently working on new cognitive domains to augment the current set of domains. These prospective cognitive domains are:

- *Speech and articulation*, fluency of speech, and comprehension of visual information
- *Eye movement*, eye focus and hand-eye coordination

**2.3. Data analysis**

During an Altoida test, the hand-held device is recording data from various sensors. This raw sensor data is difficult to interpret and difficult to compare between subjects. For the purpose of Alzheimer's prediction and cognitive domain scoring, we reduce the dimensionality of the data by means of feature extraction.

**2.4. Recorded data**

During an Altoida test we record (sensor) data from the following sources:

- ACC, accelerometer, measuring the acceleration in all three axes, relative to the device with the gravity component filtered out
- ATT, attitude meter or, gyroscope, measuring the angle of rotation over all three axes
- TOUCH, screen touches combined with an estimate of the applied pressure
- PATH, the trajectory of the device through the physical space, as estimated by the tablet based on filtering the data of the accelerometer and gyroscope.

In addition, in prospective versions of the Altoida test we will record the following additional information:

- SPEECH, speech analysis of the speech subtask. The speech is analyzed on the device and the resulting analysis is stored as raw data.
- EYES, analysis of eye focus and blinks, only stored if eye tracking is enabled.

**2.5. Feature extraction**

During the Altoida acitivity batter, the handheld device records data from all available sensors for later analysis. Depending on the available sensors in the device, this data can include accelerometer data, gyroscope data, screen touches, speech audio recording, screen brightness, ambient brightness, eye tracking, and compass data. Thus, a single session of a single subject can lead to millions of data points over all sensors. To reduce the large dimensionality of the raw sensor data, we applied various feature extraction techniques depending on the performed test. For example, in the drawing tests the subjects were asked to draw a specific shape with their finger on the touch screen. Given the raw screen touches, we extracted various features such as drawing precision, drawing speed, number of breaks in touches, etc. Likewise, during the augmented reality test we collected accelerometer and gyroscope data which lend itself to a Fourier analysis. Thus, the obtained frequency magnitudes could again be used as a feature. The extracted features were subsequently used to assess the overall cognitive performance of a subject. The performance is presented as a set of scores over several cognitive domains.
Feature extraction is a data reduction technique aimed at describing the data as a set of non-redundant and informative metrics. The sensor data described in the previous section are what we consider "raw" data. Using feature extraction, we are able to go from this raw data to interpretable features. For example, the data from the accelerometer is just a long list of measured accelerations in all three axes. By itself this does not say much about the subject. By means of feature extraction we are able to extract, for example, micro-tremors from this data. These micro-tremors are far more concise and informative than the original raw data.

**2.6. Diagnostic Criteria**

In clinical practice, the diagnosis of MCI, like AD dementia, cannot be made solely by laboratory or other tests, but requires the judgment of a clinician who takes into account clinical, cognitive, and functional criteria that define the syndrome. Moreover, clinical diagnosis in disease-modifying treatment trials is increasingly advanced to the prodromal phase of Alzheimer's disease (AD), and inclusion criteria are based on biomarkers rather than clinical symptoms, as defined by the revised National Institute on Aging (NIA) and Alzheimer's Association (AA)[7]. The diagnosis was made as per the Comprehensive Medical Follow-up (CMFV) assessments diagnostic criteria. CMFV assessments will normally be captured during a separate clinic visit to decrease site and participant burden at regular in-clinic visits; The CMFV should ideally be conducted within 30 days of the regular in-clinic visit in which the trigger(s) was noted. Assessments conducted at a CMFV included physical and neurological exam, the CDR, Neuropsychiatric Inventory Questionnaire, ADCS CGIC–MCI, blood chemistry, and tests used in evaluating cognitively impaired and dementia individuals (e.g., thyroid profile, vitamin B12). At the second of two consecutive CMFVs, a magnetic resonance imaging (MRI) scan can be performed if all other data corroborated an MCI diagnosis, to allow the investigator to rule out structural brain issues (e.g., tumor, stroke) as a potential cause of impairment. After each CMFV, considering the totality of clinical information and after consultation with the site neuropsychologist, the site investigator can assign a provisional clinical diagnosis to the participant's case from one of four diagnostic categories: reversible cognitive impairment (e.g., thyroid dysfunction); irreversible cognitive impairment (e.g., stroke, tumor, etc.); AD dementia; or non-AD dementia.

### 3. Data (Altoida and ADNI)

#### 3.1. MMSE and FAQ subitem scores

**TABLE S1. Description of item scores of MMSE**

| | MMSE Subitem Scores | Description |
|---|---|---|
| 1 | MMSE Attention Concentration | Clinical test used to assess mental function |
| 2 | MMSE Language | Tests related to naming a pencil and a watch, repeating words, and carrying out complex commands like drawing a figure measured |
| 3 | MMSE Memory Recall | Registration recall |
| 4 | MMSE Orientation | Testing orientation to time and place |
| 5 | MMSE Working Memory Registration | Testing related to repeating names prompts |

**TABLE S2. Description of item scores of FAQ**

| | FAQ Subitem Scores | Description |
|---|---|---|
| 1 | FAQFORM | assembling tax records, business affairs, or other papers, Partial score of FAQ |
| 2 | FAQBEVG | heating water, making a cup of coffee, turning off the stove. Partial Score, FAQ |
| 3 | FAQGAME | playing a game of skill such as bridge or chess, working on a hobby. Partial Score, FAQ |
| 4 | FAQFINAN | writing checks, paying bills, or balancing checkbook. Partial Score, FAQ |
| 5 | FAQMEAL | preparing a balanced meal, Partial score of FAQ |
| 6 | FAQTV | paying attention to and understanding a TV program, book, or magazine, Partial score of FAQ |
| 7 | FAQREM | remembering appointments, family occasions, holidays, medications, Partial score of FAQ |
| 8 | FAQSHOP | shopping alone for clothes, household necessities, or groceries, Partial Score of FAQ |
| 9 | FAQTRAVL | traveling out of the neighborhood, driving, or arranging to take public transportation, Partial score of FAQ |
| 10 | FAQEVENT | keeping track of current events, Partial Score of FAQ |

##### 3.2 Definition of Modules in Altoida and ADNI Data

1. Modules and their description for Altoida data are illustrated in Table S1 and for ADNI data are illustrated in Table S2. In the Altoida data, module 1 to 11 consists of measures that are the outcome of the virtual reality game. module 12 to 16 are pen and paper tests that rank the subject on a given scale of cognition. In ADNI data, module 1 represents the molecular aberrations in the brain reflected by cerebrospinal fluid (CSF), Module 2 represents the brain volume measurements, Module 3 represents the imaging measurements comprising AV45 and FDG, Module 4 to 22 represent the genetic burden scores for each molecular mechanism affected in AD, Module 23 to 27 are pen and paper tests that rank the subject on a given scale of cognition.

**TABLE S3. Modules and their description defined for Altoida data set**

| | Module name | Description |
|---|---|---|
| 1 | AR Global Telemetry Variance | The variance in telemetry (accelerometer and gyroscope) over the entire duration of the AR test |
| 2 | AR Intro Read Times | The time the subject required to read the introduction to the AR test. |
| 3 | AR Object Finding | Features pertaining to finding an object, such as time required to find the next object, distance travelled while searching, etc. |
| 4 | AR Object Placement | Features pertaining to placing an object, such as time taken to find a suitable surface, distance travelled, holding the device steady, etc. |
| 5 | AR Object Placement FFT | Fast Fourier Transform frequency spectrum analysis of a few seconds prior to placing an object. These are special enough to warrant their own group. |
| 6 | AR Place and Find Telemetry Variance | The telemetry variance in the moments before placing and finding objects. In contrast to the global telemetry variance this disregards the walking periods. |
| 7 | AR Screen Button Presses | Touch screen data of button pressed during the AR test, such as pressure, and touch accuracy. |
| 8 | BIT DOT Motor Instruction Reading Time Ratios | The difference in time when reading the instructions to the motor tests for the second time. |
| 9 | Motor Drawing Features | Features pertaining to the finger drawing tests |
| 10 | Motor Tapping Features | Features pertaining to the finger tapping tests. |
| 11 | Motor Test Durations | The total duration of each test part. |
| 12 | MMSE Attention Concentration | Clinical test used to assess mental function |
| 13 | MMSE Language | Tests related to naming a pencil and a watch, repeating words, and carrying out complex commands like drawing a figure measured |
| 14 | MMSE Memory Recall | Registration recall |
| 15 | MMSE Orientation | Testing orientation to time and place |
| 16 | MMSE Working Memory Registration | Testing related to repeating names prompts |

**TABLE S4. Modules and their description defined for ADNI data set**

| | Module name | Description |
|---|---|---|
| 1 | csf_VIS1 | Abeta, tau and ptau csf biomarkers at baseline |
| 2 | volume_VIS (1,6,12,24) | 6 brain volumes, entorhinal, hippocampus, ventricles, whole brain, middle temporal and fusiform at baseline, month 6, month 12 and month 24 (brain volumes were normalized with respect to intracranial brain volume (ICV)) |
| 3 | Imaging_VIS(1) | imaging measures such as fluorodeoxyglucose-positron emission tomography (FDG PET) and Alzbio3 kits and Florbetapir (AV45) amyloid PET at baseline |

| | | |
|---|---|---|
| 4 | ADAM_Metallopeptidase_subgraph | "rs4575098" "rs2277027" "rs1422795" "rs7174386" "rs12906705" "rs28455654" "rs383902" |
| 5 | Amyloidogenic_subgraph | "rs17571" "rs676134" "rs2829946" "rs2830088") |
| 6 | APOE_subgraph | "rs405509" "rs439401" |
| 7 | Apoptosis_signaling_subgraph | "rs1136410" "rs1805411" "rs1469926" "rs319724" "rs827423" "rs6902771" "rs8006145" "rs10144225" "rs10137185" "rs2667543") |
| 8 | ATP_binding_cassette_transport_subgraph | "rs1045642" "rs6949448" "rs2235046" "rs1128503" "rs10276036" "rs1202169" "rs1202168" "rs1202167" "rs1883023" "rs2777802" "rs3818689" "rs2066715" "rs2066718" "rs4149308" "rs2066717" "rs4149303" "rs4149301" "rs2297399" "rs2297400" "rs3824479" "rs2472384" "rs2253304" "rs2253182" "rs2253175" "rs2253174" "rs2253172" "rs2230806" "rs2243313" "rs2482420" "rs2487058" "rs2487059" "rs2230805" "rs3847300" "rs3847303" "rs3905000" "rs2575876" "rs12826" "rs7067971" "rs11190305" "rs1283816" "rs1283817" "rs829079" "rs1283822" "rs3752229" "rs3752232" "rs3764650" "rs3752240" "rs3752242" "rs2279796" "rs4147932" |
| 9 | Axonal_guidance_subgraph | "rs1354269" "rs12364788" "rs17614100" "rs7112354" |
| 10 | Caspase_subgraph | "rs2027432" "rs10159239" "rs12130711" "rs1143634" "rs2276575" "rs13430599" "rs10194375" "rs13426725" "rs17014923" "rs6743470" "rs4663098" "rs7561528" "rs744373" |
| 11 | Chemokine_signaling_subgraph | "rs1024611" "rs991804" |
| 12 | Cholesterol_metabolism_subgraph | "rs5174" "rs3737983" "rs2297663" "rs2297660" "rs3820198" "rs7551288" "rs9371201" "rs1799986" "rs12435918" |
| 13 | GSK3_subgraph | "rs2873950" "rs3108749" "rs6438552" |
| 14 | Inflammatory_response_subgraph | "rs2243248" "rs2243290" "rs7748777" "rs7759295" "rs7072793" "rs6074022" "rs1569723" "rs6032678" |
| 15 | Insulin_signal_transduction | “rs1999763” |
| 16 | Interferon_signaling_subgraph | "rs1554606" "rs8038734" |
| 17 | Interleukin_signaling_subgraph | "rs4537545" "rs4129267" "rs4240872" "rs7514452" "rs1800896" "rs4848300" "rs17561""rs4848304" "rs1143634" "rs2243248" "rs2243290" "rs1554606" "rs10975516" "rs1330383""rs10815398" "rs7072793" "rs4072111" "rs11857713" "rs4778636" "rs7197333" |
| 18 | Lipid_metabolism_subgraph | "rs2228467" "rs6444175" "rs11742194" "rs3846662" "rs5909" "rs12435918" "rs5882" |
| 19 | Matrix_metalloproteinase_subgraph | "rs10836653" "rs4382897" "rs17337649" "rs645419" "rs2241715" |
| 20 | Nerve_growth_factor_subgraph | “rs3775256” |
| 21 | Tau_protein_subgraph | "rs242557" "rs3785883" |
| 22 | Wnt_signaling_subgraph | "rs2873950" "rs3108749" "rs6438552" "rs29645" "rs7901695" "rs7903146" |
| 23 | MMSE Language_VIS (1,6,12, 24, 36) | Tests related to naming a pencil and a watch, repeating words, and carrying out complex commands like drawing a figure measured at baseline, month 6, 12 , 24 and 36.<br>Ranges between 0 and 5. |
| 24 | MMSE Memory Recall_VIS (1,6,12, 24,36) | Registration recall measured at baseline, month 6, 12, 24 and 36<br>Ranges between 0 and 5. Ranges between 0 and 3. |

| | | |
|---|---|---|
| 25 | MMSE Orientation_VIS (1,6,12, 24, 36) | Testing orientation to time and place at baseline, month 6, 12, 24 and 36. Ranges between 0 and 10. |
| 26 | MMSE Working Memory Registration_VIS (1,6,12, 24, 36) | Testing related to repeating names prompts at baseline, month 6, 12, 24 and 36. Ranges between 0 and 3. |
| 27 | MMSE_Attention_Concentration_VIS (1,6,12, 24, 36) | Clinical test used to assess mental function at baseline, month 6, 12, 24 and 36. Ranges between 0 and 5. |

### 4. Details on VAMBN training

**4.1 The VAMBN workflow follows the following steps:**

1. Definition of modules that summarizes the original input features (Table S1 and S2)
2. Encoding of modules into lower dimensional latent distributions (multivariate gaussians) via a variational autoencoder for heterogeneous and incomplete data (HI-VAE). The training procedure of HI-VAEs for each module included a hyperparameter optimization over the following parameters:

- Learning rate ∈ {0.01, 0.001]
- Mini batch size ∈ {16, 32}
- Weight Decay ∈ {0, 0.001, 0.01}

  The 3-fold cross-validated reconstruction loss was used as an objective function to evaluate each candidate parameter set

3. Structure and Parameter Learning

   Structure Learning: Structure learning of a modular Bayesian network (MBN) between the encoded modules: As most edges in the MBN structure are not known, therefore we need to deduce them from the data. However, MBN structure learning is an NP hard problem, the number of possible DAGs grow super-exponentially with the number of nodes (Chickering et al., 2004). Hence, the search space of possible network structures should a priori be restricted as much as possible. We follow two essential strategies for this purpose:

   a) Variables were grouped in the raw data into autoencoded modules, as explained above.
   b) Causal constraints as prior knowledge was imposed while learning the augmented BN connected modules in order to restrict the search space and to allow correct causal orientation of as many edges as possible.

   After structure learning, parameters of the MBN were fitted. This was done via maximum likelihood. That means for each Gaussian node a linear regression was fitted, in which the parents of the node were used as predictors. For each discrete node parameters were determined via conditional probability tables.

**4.2 Causal constraints for VAMBN training on Altoida Data:**

During structure learning the following constraints were imposed:

- Demographic features such as age and gender cannot be influenced by any other feature or by each other.
- Amyloid status of a subject cannot be affected by any other feature.
- Cognitive features and digital biomarkers cannot affect clinical diagnosis (it can be only affected by Amyloid)
- No other feature can affect education except age and gender
- An auxiliary variable representing the missingness of a certain feature can only influence that particular feature at a later visit.

**4.3 Causal constraints for VAMBN training on ADNI Data:**

During structure learning the following constraints were imposed:

- Demographic features such as age and gender cannot be influenced by any other feature or by each other.
- Modules of brain volumes can be related to each other, but they cannot be influenced by modules of MMSE or FAQ features.
- Longitudinal measures must follow the right temporal order, i.e., there are no edges pointing backwards in time.
- Modules of genetic burden scores can be related to each other, but they cannot be influenced by other modules except the demographics.
- The digital features can only influence each other and no other feature or module.
- Diagnostic status cannot influence any other feature except the digital features

- Auxiliary and missing visit nodes were connected to their respective counterparts at the next time point, accounting for a correlation between these measures over time, e.g., through study dropout.

Structure learning was conducted via greedy hill climbing (hc)[8]. Hill climbing explores the directed acyclic search space by single arc direction, removals and reversals and it avoids local optima with random restarts. Score based algorithms have been empirically observed to show a more robust behaviour in terms of network reconstruction accuracy than constraint-based methods for mixed type of data (both discrete and continuous, specifically for smaller sample sizes).

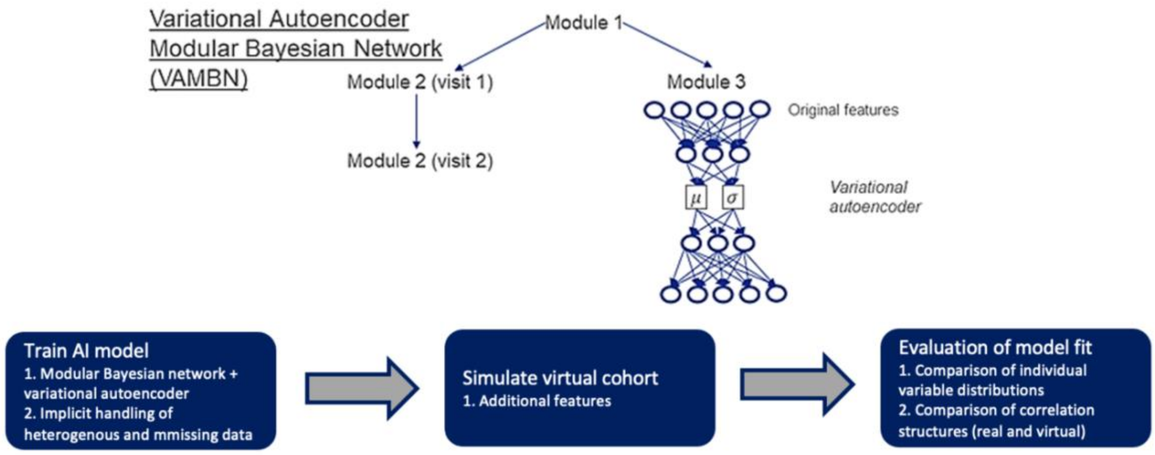

FIGURE S1. Workflow describing the steps followed in VAMBN

#### 5. Predicting digital measures (digital tasks and digital cognitive domains) in ADNI

##### 5.1 Evaluating the predictability of digital measures and cognitive domains

Prediction performance was evaluated via the normalized root mean squared error (NRMSE):

$$\mathrm{NRMSE} = \frac{1}{Y_{\max} - Y_{min}} \sqrt{\frac{\sum_{i=1}^{N} (Y_i - \hat{Y}_i)^2}{N}}$$

Here $Y_i$ and $\widehat{Y_i}$ denote the real and inferred DM, respectively. Bayesian inference was performed via the likelihood weighting approach implemented in R-package bnlearn[9]. Notably the approach allows for making predictions in a patient, in which no DM has been observed. This amounts to fixing the value of all nodes in the BN (except the DMs) to their observed values in the patient and subsequently running inference. Accordingly, it is also possible to predict DMs in ADNI based on a VAMBN model learned from Altoida data: For each ADNI patient we fixed the value of all nodes representing demographic features, diagnostic status and MMSE scores in the BN and then inferred the most likely value of DMs. This procedure was run for each visit in ADNI.

10-fold cross-validation errors of VAMBN (more precisely the augmented BN) were compared against a standard Random Forest (RF) regression model using 100 regression trees. The following additional hyper-parameters of the RF were considered:

a. mtry = {2, 3, 4}
b. splitrule = {"variance", "extratrees","maxstat"}
c. min. node. size = {10, 20}

These hyper-parameters were tuned via a grid search, in which each hyper-parameter combination was evaluated via an inner 10-fold cross-validation. That means there was a repeated, nested cross-validation procedure. Results are shown in Table S3 (digital tasks) and Table S4 (digital cognitive domains).
For the sake of completeness, we also evaluated, in how far MMSE subitem scores could be inferred from DMs, demographic data and diagnostic status. Corresponding results are shown in Table S5.

**TABLE S5. Prediction error (normalized root mean squared error) for digital tasks using VAMBN and a Random Forest (RF) regression model. The results shown were obtained via 10 times repeated 10-fold cross-validation procedure, and the mean and standard error (SE) is of the 10-fold cross-validated error presented.**

| Digital Task | NRMSE: Mean±SE (VAMBN) | NRMSE: Mean±SE (RF) |
|---|---|---|
| ARObjectFinding | 0.1747 ±0.0017 | 0.1611±0.0098 |
| ARScreenButtonPresses | 0.1287±0.0012 | 0.1318±0.0080 |
| MotorTestDurations | 0.1858±0.0040 | 0.2009±0.0186 |
| ARObjectPlacementFFT | 0.1647±0.0017 | 0.3532±0.0398 |
| ARPlaceAndFindTelemetryVariance | 0.2084±0.0015 | 0.1678±0.0208 |
| ARGlobalTelemetryVariance | 0.2208±0.0023 | 0.2069±0.0140 |
| ARObjectPlacement | 0.1450±0.0017 | 0.2088±0.0197 |
| BITDOTMotorInstructionReadingTimeRatios | 0.1827±0.0020 | 0.1539±0.0136 |
| MotorTappingFeatures | 0.2111±0.0046 | 0.1835±0.0266 |

**TABLE S6. Prediction error (normalized root mean squared error) for cognitive domains using VAMBN and a Random Forest (RF) regression model. The results shown were obtained via 10 times repeated 10-fold cross-validation procedure, and the mean and standard error (SE) is of the 10-fold cross-validated error presented.**

| Cognitive Domain | NRMSE: Mean±SE(VAMBN) | NRMSE: Mean±SE (RF) |
|---|---|---|
| PerceptualMotorCoordination | 0.1911±0.0024 | 0.9881±0.0881 |
| ComplexAttention | 0.1859±0.0016 | 1.2831±0.0493 |
| CognitiveProcessingSpeed | 0.1867±0.0022 | 1.4688±0.0951 |
| Inhibition | 0.1969±0.0022 | 2.366±0.1985 |
| Flexibility | 0.2379±0.0031 | 2.5013±0.1902 |
| VisualPerception | 0.2130±0.0025 | 1.8686±0.0779 |
| Planning | 0.1633±0.0015 | 1.1609±0.0924 |
| ProspectiveMemory | 0.2379±0.0024 | 1.2948±0.0888 |
| SpatialMemory | 0.1934±0.0021 | 1.5476±0.1267 |

**TABLE S7. Prediction error (normalized root mean squared error) for MMSE subscores using VAMBN and a Random Forest (RF) regression model. The results shown were obtained via a 10-fold cross-validation procedure, and the mean and standard error (SE) of the 10-fold cross-validated error is presented.**

| MMSE Subitem | NRMSE: Mean±SE(VAMBN) | NRMSE: Mean±SE (RF) |
|---|---|---|
| MMSE Attention Concentration | 0.4149±0.0011 | 1.4179±0.0663 |
| MMSE Language | 0.3481±0.0008 | 1.0407±0.0843 |
| MMSE Memory Recall | 0.4038±0.0015 | 0.6920±0.0490 |
| MMSE Orientation | 0.3718±0.0008 | 1.6934±0.1067 |
| MMSE Working Memory Registration | 0.1298±0.0003 | 0.5263±0.4468 |

##### 5.2 Predicting the DMs in ADNI using common measures from Altoida

The VAMBN model trained on Altoida was used to predict the DMs in ADNI data. The measures observed in both data sets which includes diagnosis, demographics (age, education, gender), MMSE subitems), and tested for the predictability of DMs in section 5 in were used as the predictor variables. We used the bayes likelihood (bayes-lw) [9] methods from the bnlearn package for this purpose where we average the likelihood weighting simulations using all available nodes as evidence (here we take the common features mention above, except node that is being predicted) to compute the predicted values. The value of the predicted variable is the expected value of the conditional distribution. We compute the predicted DMs at each time point (baseline, month 6, 12, 24 and 36) as we have the values of the common features available at each time point.

#### 6. Observed correlations between variables

##### 6.1. Altoida

The scatter plots in Figure S2 depict correlations between variables, which were found connected according to VAMBN. The figures also display the spearman rank correlation coefficient (R), adjusted confidence intervals (95%) and multiple testing adjusted p values (p. Adj). Multiple testing correction was done using Holm adjustment.

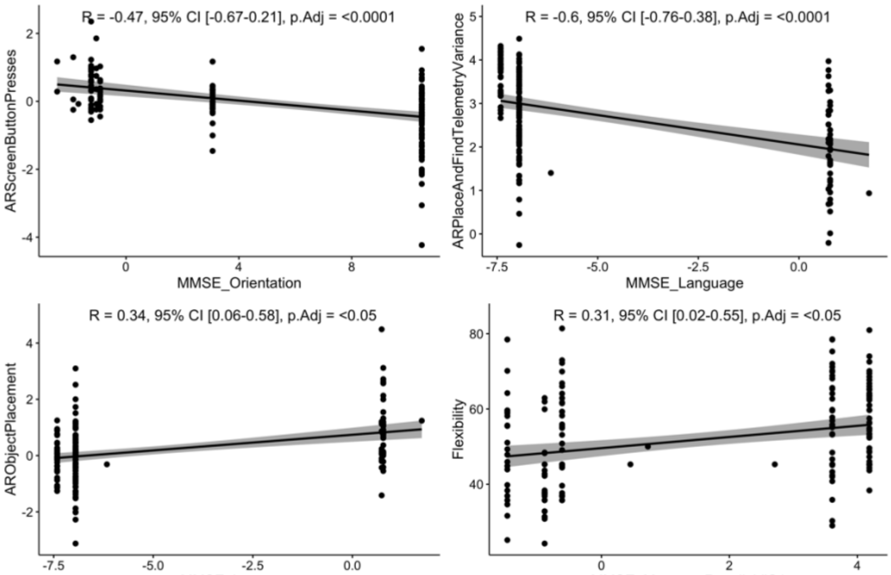

FIGURE S2. Scatter plot between different pairs of variables in Altoida data: MMSE Orientation and AR Screen Button Presses (top left), MMSE Language and AR Place and Find Telemetry Variance (top right), MMSE Language and AR Object Placement (bottom left)) and MMSE Memory Recall Registration and Flexibility (bottom right).

#### 6.2. ADNI

The scatter plots in Figure S3, S4, S5 depict correlations between variables, which were found connected according to VAMBN. The figures also display the spearman rank correlation coefficient (R) adjusted confidence intervals (95%) and multiple testing adjusted p values (p. Adj). Multiple testing correction was done using Holm adjustment. Figure S3 depicts the connections that were commonly found in VAMBN networks learned from ADNI and Altoida data, whereas Figure S4 and S5 illustrated connections that were newly observed in ADNI.

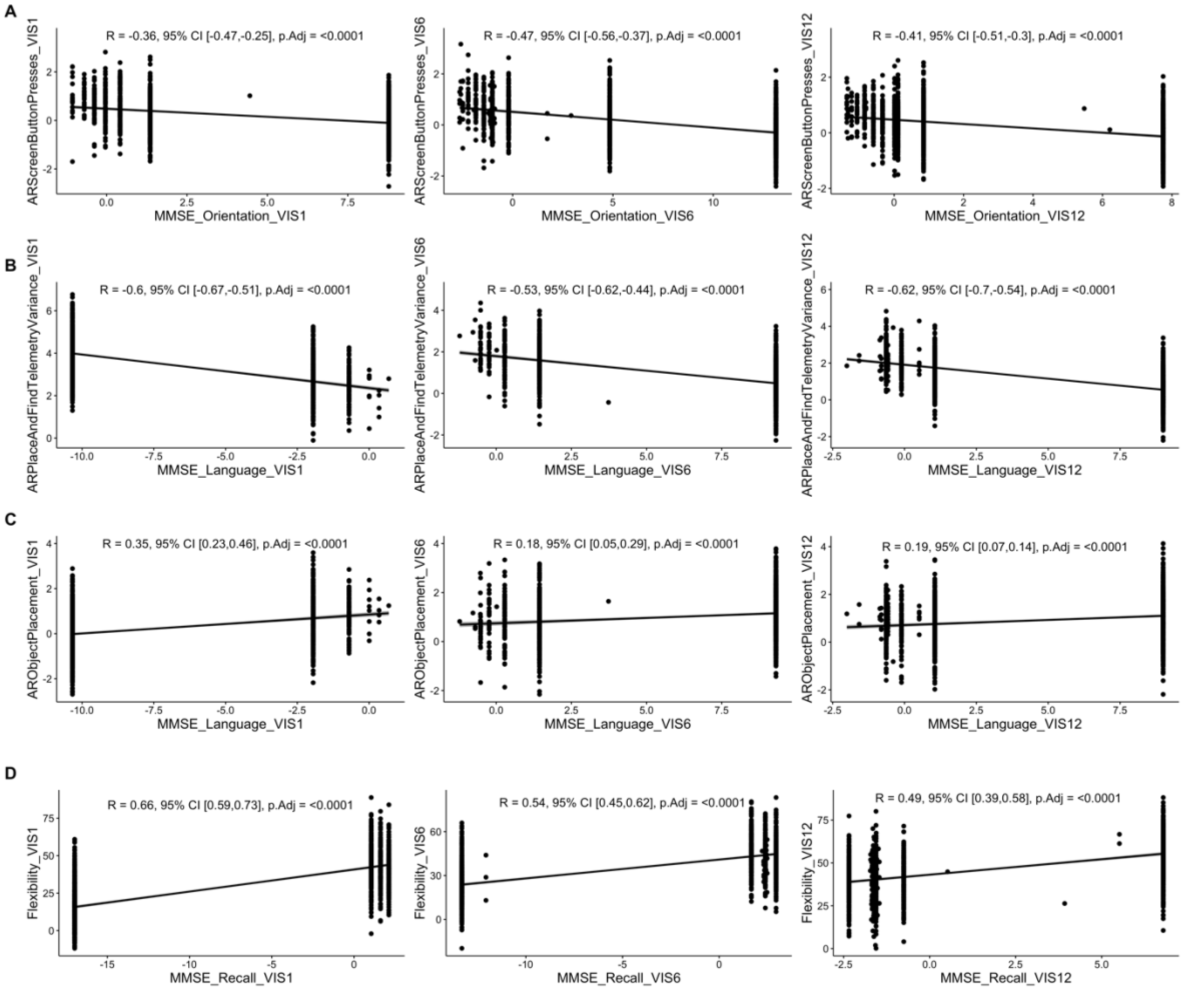

FIGURE S3: Scatter plot between different pairs of variables in ADNI data: A) MMSE Orientation and AR Screen Button Presses (baseline, month 6 and month 12), B) MMSE Language and AR Place and Find Telemetry Variance (baseline, month 6 and month 12), C) MMSE Language and AR Object Placement (baseline, month 6 and month 12), D) MMSE Memory Recall Registration and Flexibility (baseline, month 6 and month 12).

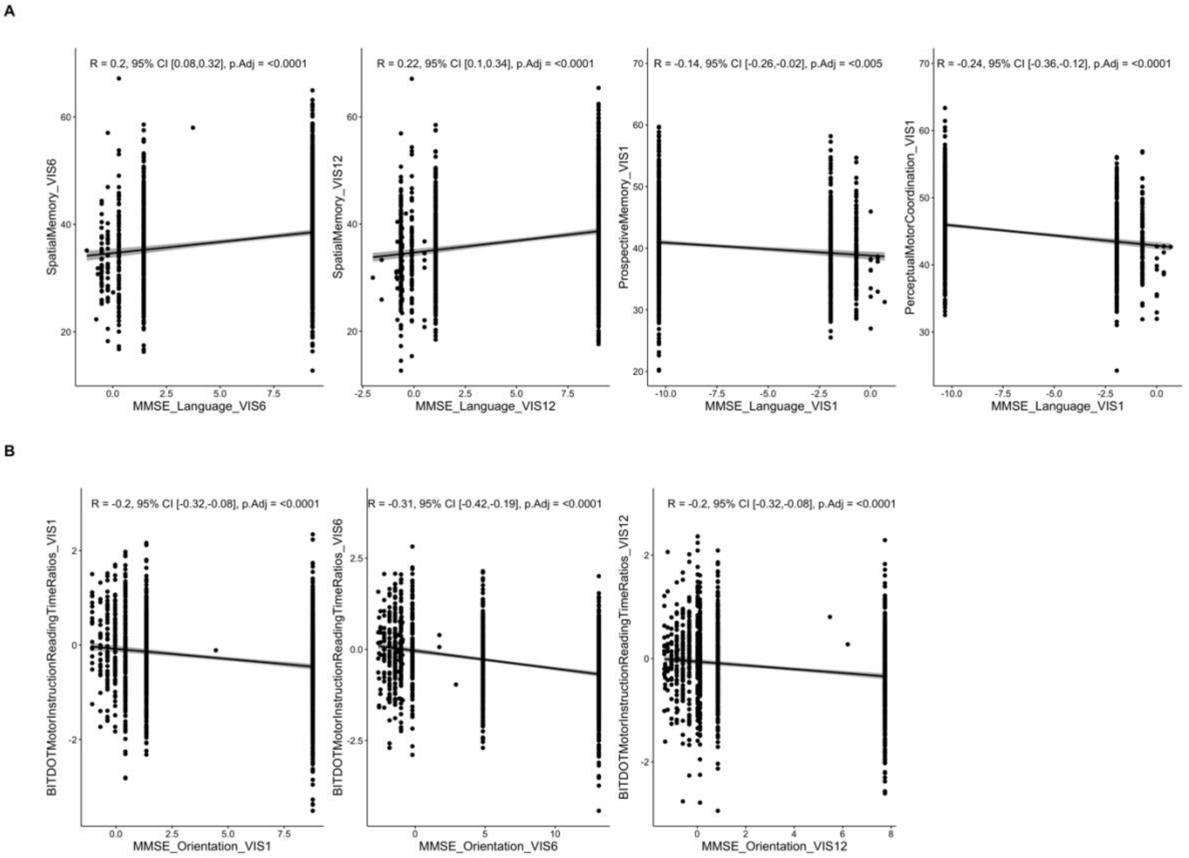

FIGURE S4: Scatter plot between different pairs of variables in ADNI data: A) MMSE Language with Spatial Memory (month 6 and 12), with Prospective Memory (at baseline) and with Perceptual Motor Coordination (at baseline) B) MMSE Orientation and AR Screen Button Presses (baseline, month 6 and month 12), FAQREM at month 24 to Motor Test Durations at month 36 and FAQFINAN at visit 12 to AR Object Finding at month 24.

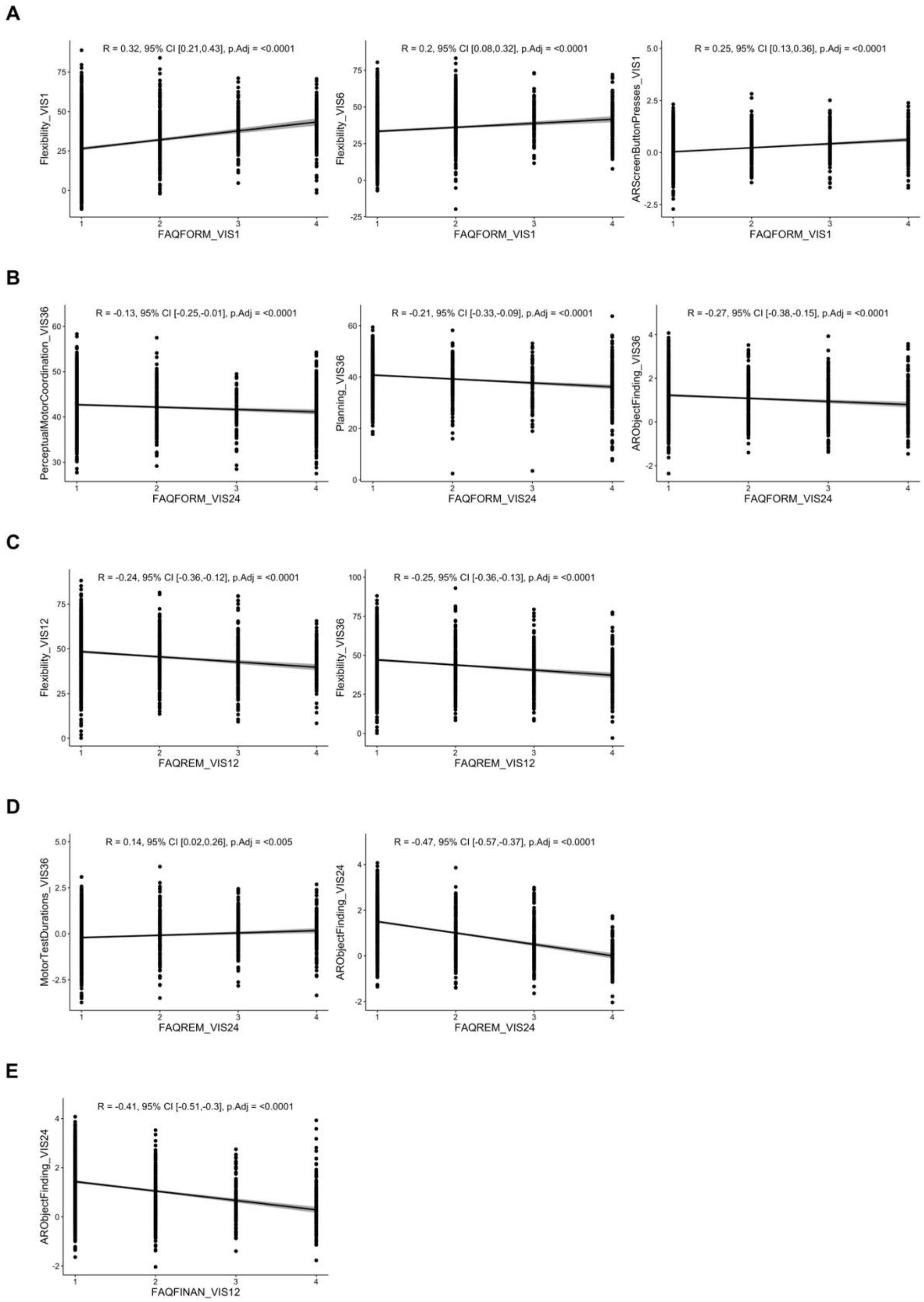

FIGURE S5: Scatter plot between different pairs of variables in ADNI data: A) FAQFORM at visit 1 to Flexibility at baseline, at month 6 and AR screen button presses at baseline B) FAQFORM at month 24 to perceptual motor coordination at month 36, planning at month 36 and AR Object Finding at month 36. C) FAQREM at month 12, to Flexibility at month 13 and month 36 D) FAQREM at month 24 to Motor Test Durations at month 36 and AR Object Finding at month 24 E) FAQFINAN at month 12 to AR object finding at month 24

#### 7. Differences in digital measures and MMSE across diagnostic states

The dependencies from digital cognitive domains, digital tasks and MMSE on the diagnostic status (CN and MCI) in Altoida are displayed in Table S8, Table S9 and Table S10 respectively. These dependencies were verified by conducting Mann Whitney U-tests.

**TABLE S8. Significance of the differences in each cognitive domain across different stages (Altoida data). Multiple testing was performed via Holm's correction.**

| Digital cognitive domains | p. adjusted (CN and MCI) | p. adjusted (MCI and Prodromal AD) | p. adjusted (MCI and Dementia) |
|---|---|---|---|
| Perceptual Motor Coordination | 0.0948 | 0.8142 | 0.0010 |
| Complex Attention | 0.1287 | 0.9474 | 0.0010 |
| Cognitive Processing Speed | <0.0001 | 0.4170 | 0.2194 |
| Inhibition | 0.7541 | 0.9474 | 1.0000 |
| Flexibility | 0.7541 | 0.0002 | 0.0653 |
| Visual Perception | 0.3215 | 0.1581 | 1.0000 |
| Planning | 0.1287 | 0.0320 | <0.0001 |
| Prospective Memory | <0.0001 | 0.0320 | 0.4901 |
| Spatial Memory | <0.0001 | 0.0173 | 0.4901 |

**TABLE S9. Significance of the differences in each digital task across different stages (Altoida data). Multiple testing was performed via Holm's correction.**

| Digital tasks | p. adjusted (CN and MCI) | p. adjusted (MCI and Prodromal AD) | p. adjusted (MCI and Dementia) |
|---|---|---|---|
| Feature 133, ARObjectFinding | <0.0001 | 0.3172 | 0.8462 |
| Feature 1037, ARObjectPlacement | 0.0106 | 0.1993 | 0.0062 |
| Feature 978, ARObjectPlacement | 0.0017 | <0.0001 | <0.0001 |
| Feature 977, ARObjectPlacement | 0.0175 | <0.0001 | <0.0001 |
| Feature 138, ARObjectFinding | <0.0001 | 1.0000 | 0.3373 |
| Feature 135, ARObjectFinding | <0.0001 | 1.0000 | 0.0041 |
| Feature 140, ARObjectFinding | 0.0106 | 1.0000 | 0.9019 |

| | | | |
|---|---|---|---|
| Feature 281, MotorTestDurations | 0.0001 | 1.0000 | 0.9019 |
| Feature 296, MotorTestDurations | <0.0001 | 0.3210 | 0.9019 |
| Feature 276, MotorTestDurations | <0.0001 | <0.0001 | 0.0001 |
| Feature 291, MotorTestDurations | <0.0001 | 0.0197 | 0.8462 |
| Feature 134, ARObjectFinding | 0.0006 | 0.3210 | 0.1157 |
| Feature 306, MotorTestDurations | <0.0001 | <0.0001 | 0.0007 |
| Feature 136, ARObjectFinding | <0.0001 | 1.0000 | 0.3373 |
| Feature 127, ARObjectFinding | 0.7919 | 0.0545 | 0.0001 |

**TABLE S10. Significance of the differences in each MMSE subitem score across different stages (Altoida data). Multiple testing was performed via Holm's correction.**

| MMSE subitem scores | p. adjusted (CN and MCI) | p. adjusted (MCI and Prodromal AD) | p. adjusted (MCI and Dementia) |
|---|---|---|---|
| MMSE_Attention_Concentration | <0.0001 | 0.6577 | 0.0430 |
| MMSE_Language | <0.0001 | 0.2197 | 0.0430 |
| MMSE_Memory_Recall | <0.0001 | 1.0000 | 0.3485 |
| MMSE_Orientation | <0.0001 | 1.0000 | 0.0224 |
| MMSE_Working_Memory_Registration | 0.2189 | 1.0000 | 0.0301 |

We also calculated the spearman rank correlation between the digital nodes having conditionally dependency on Age in Altoida (Table S10) and ADNI (Table S11).

**TABLE S11. Spearman Rank Correlation with adjusted p values (Holm's method) for digital tasks and age in Altoida**

| Demographic | Digital Measure (DM) | Spearman Rank Correlation Coefficient ($\rho$) | p-adjusted | lower.ci | upper.ci |
|---|---|---|---|---|---|
| AGE | ARScreenButtonPresses_VIS1 | 0.38 | <0.0001 | 0.10 | 0.60 |
| AGE | ARGlobalTelemetryVariance_VIS1 | -0.40 | <0.0001 | -0.62 | -0.13 |

| | | | | | |
|---|---|---|---|---|---|
| AGE | ARObjectPlacementFFT_VIS1 | 0.32 | <0.05 | 0.03 | 0.56 |

**TABLE S12. Spearman Rank Correlation with adjusted p values (Holm's method) for digital tasks and age in ADNI**

| **Diagnosis/Age** | **Digital Measure (DM)** | **Spearman Rank Correlation Coefficient ($\rho$)** | **p-adjusted** | **lower.ci** | **upper.ci** |
|---|---|---|---|---|---|
| AGE | ARGlobalTelemetryVariance_VIS1 | -0.39 | <0.0001 | -0.49 | -0.28 |
| AGE | BITDOTMotorInstructionReadingTimeRatios_VIS1 | -0.14 | <0.05 | -0.26 | -0.02 |
| AGE | ARScreenButtonPresses_VIS1 | 0.27 | <0.0001 | 0.15 | 0.38 |
| AGE | ARObjectPlacementFFT_VIS1 | 0.23 | <0.0001 | 0.11 | 0.34 |
| AGE | ARGlobalTelemetryVariance_VIS6 | -0.36 | <0.0001 | -0.47 | -0.25 |
| AGE | ARScreenButtonPresses_VIS6 | 0.26 | <0.0001 | 0.14 | 0.37 |
| AGE | ARObjectPlacementFFT_VIS6 | 0.18 | <0.0001 | 0.05 | 0.30 |
| AGE | ARGlobalTelemetryVariance_VIS12 | -0.40 | <0.0001 | -0.50 | -0.29 |
| AGE | BITDOTMotorInstructionReadingTimeRatios_VIS12 | -0.22 | <0.0001 | -0.33 | -0.09 |
| AGE | ARScreenButtonPresses_VIS12 | 0.26 | <0.0001 | 0.14 | 0.37 |
| AGE | ARObjectPlacementFFT_VIS12 | 0.21 | <0.0001 | 0.09 | 0.33 |
| AGE | Inhibition_VIS1 | -0.17 | <0.0001 | -0.28 | -0.04 |

#### 8. VAMBN model trained on common Altoida / ADNI Features

We selected the common features found in both ADNI and Altoida and then trained the model on ADNI data on these common features. The network of common features is illustrated in Figure S6.

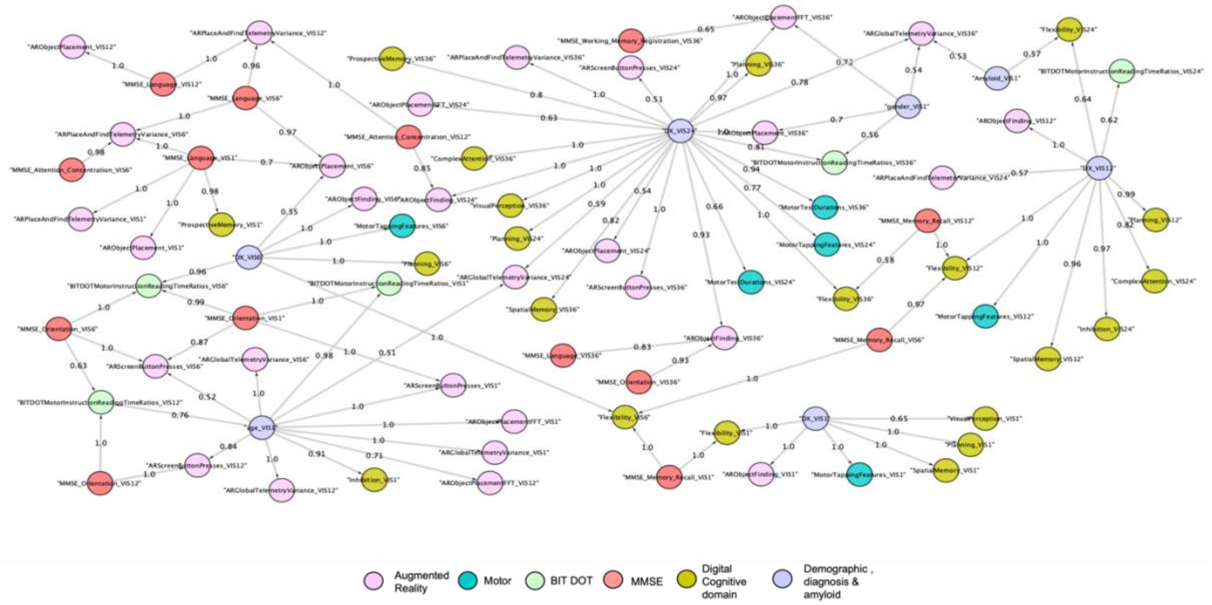

FIGURE S6. Variable dependencies identified in the common measure of ADNI dataset with Altoida dataset in 1000 bootstrapped Bayesian Network (BN) (strength >=0.5 and direction >= 0.5) reconstructions. These edges indicate variable dependencies that are found commonly in bootstrapped BN reconstruction, the edges from MMSE, diagnosis and age directly linked to the DMs were selected for visualisaton.

#### 9. Evaluation of the fit of VAMBN models

To evaluate the quality of the fit of our VAMBN models we took advantage of the fact that VAMBN is a generative model. Therefore, we can use a trained VAMBN model to generate synthetic data. The closer the synthetic data is to the real data, the better the fit of the model. In agreement to our earlier publication[10] we generated as many synthetic subjects as real patients and mathematically assessed the model fit in two ways: a) agreement of the marginal distributions of individual variables; b) agreement of the learned correlation structure. More specifically, for a) we used the Kullback-Leibler divergence as a mathematical measure to quantify the difference. For b) we calculated the Frobenius norm of the difference between the real Pearson correlation matrix and the synthetic one. This quantity was divided by the Frobenius norm of the real correlation matrix, yielding a relative error.
The number of these plots is very large. We thus only show selected results here. The distributions for selected variables in the Altoida data are depicted in Figures S7, S8 and S9. Heatmaps depicting correlation matrices and the distribution of correlation coefficients are shown in Figure S10. The distribution plots for selected variables in the ADNI data are shown in Figures S11 and S12. Heatmaps depicting the correlation matrices and the distribution of correlation coefficients are shown in Figure S13. Plots and tables of the remaining variables can be found under https://gitlab.scai.fraunhofer.de/meemansa.sood/digiad.

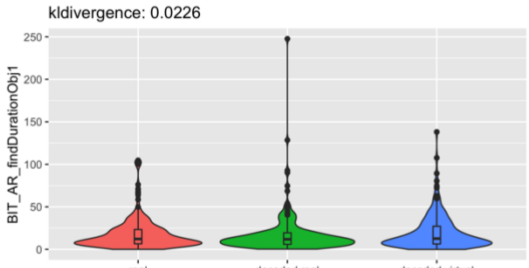

| Type | Mean | SD | 25% | Median | 75% |
|---|---|---|---|---|---|
| real | 18.28 | 18.93 | 6.48 | 12.14 | 23.32 |
| decoded real | 17.57 | 24.48 | 5.89 | 11.81 | 19.43 |
| decoded virtual | 19.84 | 20.53 | 6.28 | 12.61 | 27.16 |

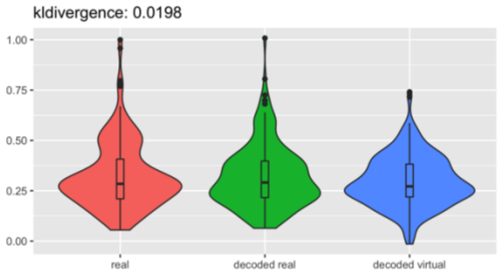

| Type | Mean | SD | 25% | Median | 75% |
|---|---|---|---|---|---|
| real | 0.32 | 0.16 | 0.21 | 0.28 | 0.41 |
| decoded real | 0.32 | 0.16 | 0.22 | 0.29 | 0.4 |
| decoded virtual | 0.3 | 0.13 | 0.22 | 0.27 | 0.38 |

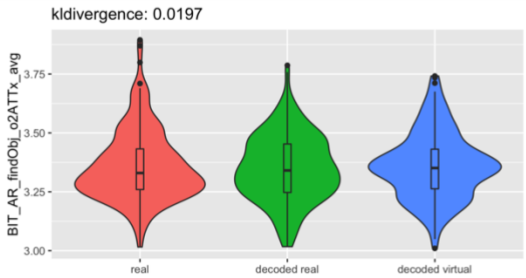

| Type | Mean | SD | 25% | Median | 75% |
|---|---|---|---|---|---|
| real | 3.36 | 0.15 | 3.26 | 3.33 | 3.43 |
| decoded real | 3.35 | 0.14 | 3.25 | 3.34 | 3.45 |
| decoded virtual | 3.36 | 0.14 | 3.26 | 3.35 | 3.43 |

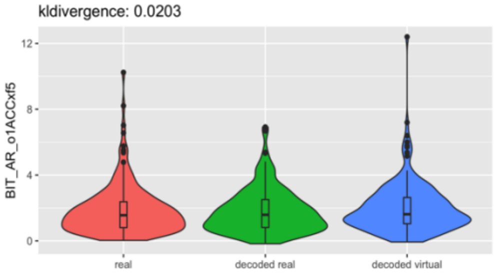

| Type | Mean | SD | 25% | Median | 75% |
|---|---|---|---|---|---|
| real | 1.89 | 1.52 | 0.81 | 1.56 | 2.38 |
| decoded real | 1.81 | 1.34 | 0.81 | 1.59 | 2.51 |
| decoded virtual | 2 | 1.53 | 1.04 | 1.62 | 2.64 |

FIGURE S7. Examples of real and simulated subjects generated via the VAMBN model trained on Altoida data. The Figure compares the marginal distributions of selected variables related to Back in Time (BIT) task for real patients (red), virtual / simulated subjects (blue) and real patients decoded via the HI-VAE model (green) Tables show summary statistics of the distributions (mean, standard deviation, 25% quantile, median, 75% quantile). KL-divergence between the decoded real and decoded virtual patients is mentioned on the top of each plot.

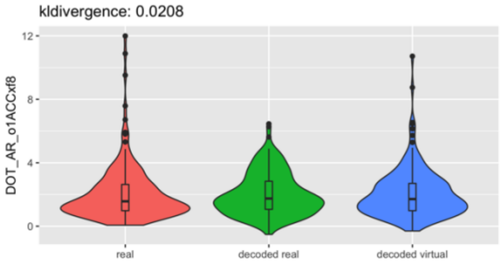

| Type | Mean | SD | 25% | Median | 75% |
|---|---|---|---|---|---|
| real | 2.07 | 1.77 | 0.98 | 1.57 | 2.63 |
| decoded real | 2.02 | 1.33 | 1.06 | 1.75 | 2.84 |
| decoded virtual | 2.06 | 1.54 | 0.98 | 1.71 | 2.7 |

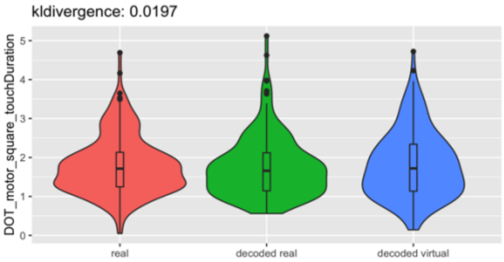

| Type | Mean | SD | 25% | Median | 75% |
|---|---|---|---|---|---|
| real | 1.8 | 0.75 | 1.25 | 1.71 | 2.13 |
| decoded real | 1.76 | 0.78 | 1.14 | 1.66 | 2.12 |
| decoded virtual | 1.82 | 0.86 | 1.13 | 1.72 | 2.34 |

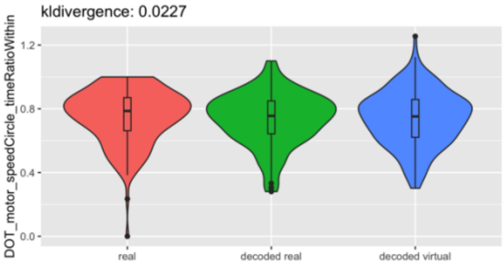

| Type | Mean | SD | 25% | Median | 75% |
|---|---|---|---|---|---|
| real | 0.75 | 0.17 | 0.66 | 0.79 | 0.87 |
| decoded real | 0.74 | 0.16 | 0.64 | 0.76 | 0.85 |
| decoded virtual | 0.74 | 0.17 | 0.62 | 0.75 | 0.86 |

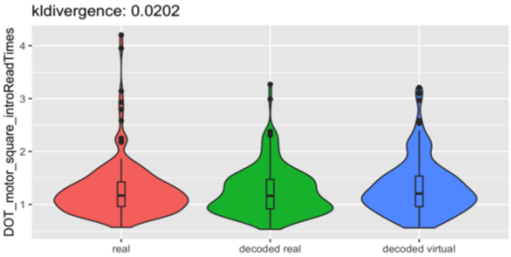

| Type | Mean | SD | 25% | Median | 75% |
|---|---|---|---|---|---|
| real | 1.27 | 0.52 | 0.96 | 1.17 | 1.43 |
| decoded real | 1.25 | 0.45 | 0.92 | 1.16 | 1.47 |
| decoded virtual | 1.31 | 0.51 | 0.96 | 1.21 | 1.54 |

FIGURE S8. Examples of real and simulated subjects generated via the VAMBN model trained on Altoida data. The Figure compares the marginal distributions of selected variables related to Day out Tasks (DOT) tasks for real patients (red), virtual / simulated subjects (blue) and real patients decoded via the HI-VAE model (green) Tables show summary statistics of the distributions (mean, standard deviation, 25% quantile, median, 75% quantile). KL-divergence between the decoded real and decoded virtual patients is mentioned on the top of each plot.

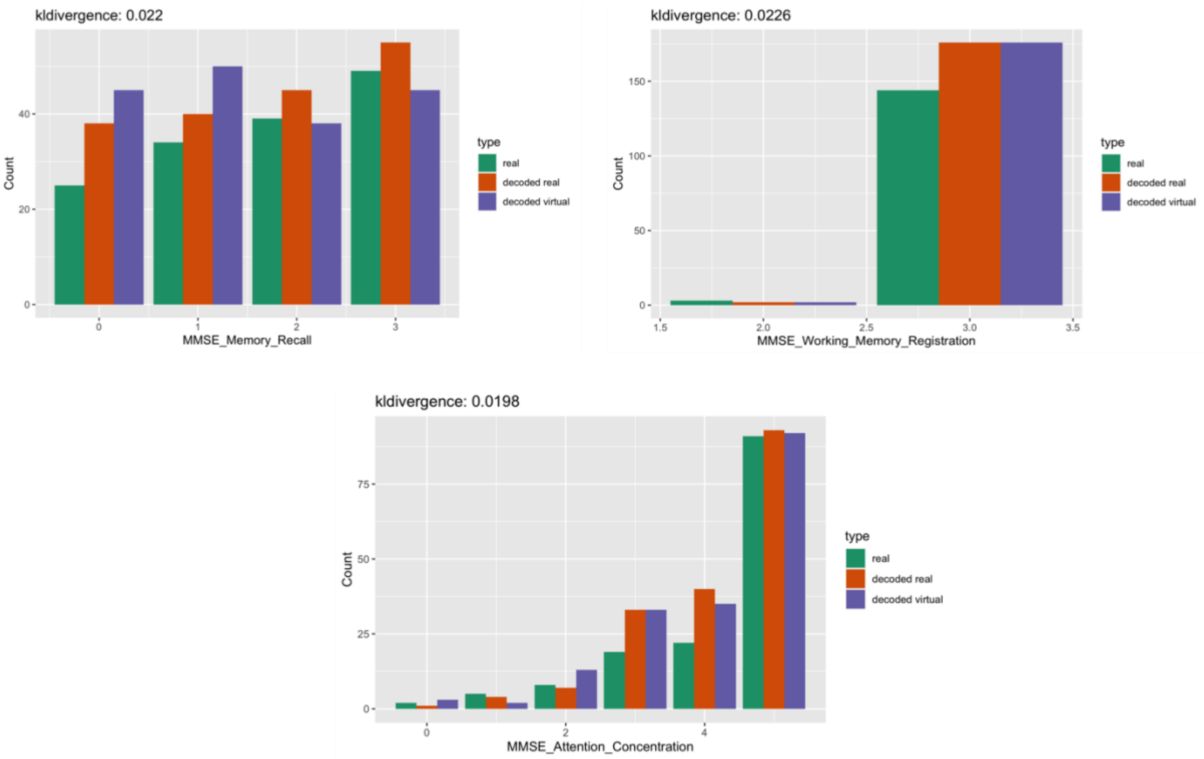

FIGURE S9. Examples of real and simulated subjects generated via the VAMBN model trained on Altoida data The Figure compares the distributions of variables related to MMSE scores for real patients (green), virtual / simulated patients (purple) and real patients decoded via the HI-VAE model (red). KL-divergence between the decoded real and decoded virtual patients is mentioned on the top of each plot.

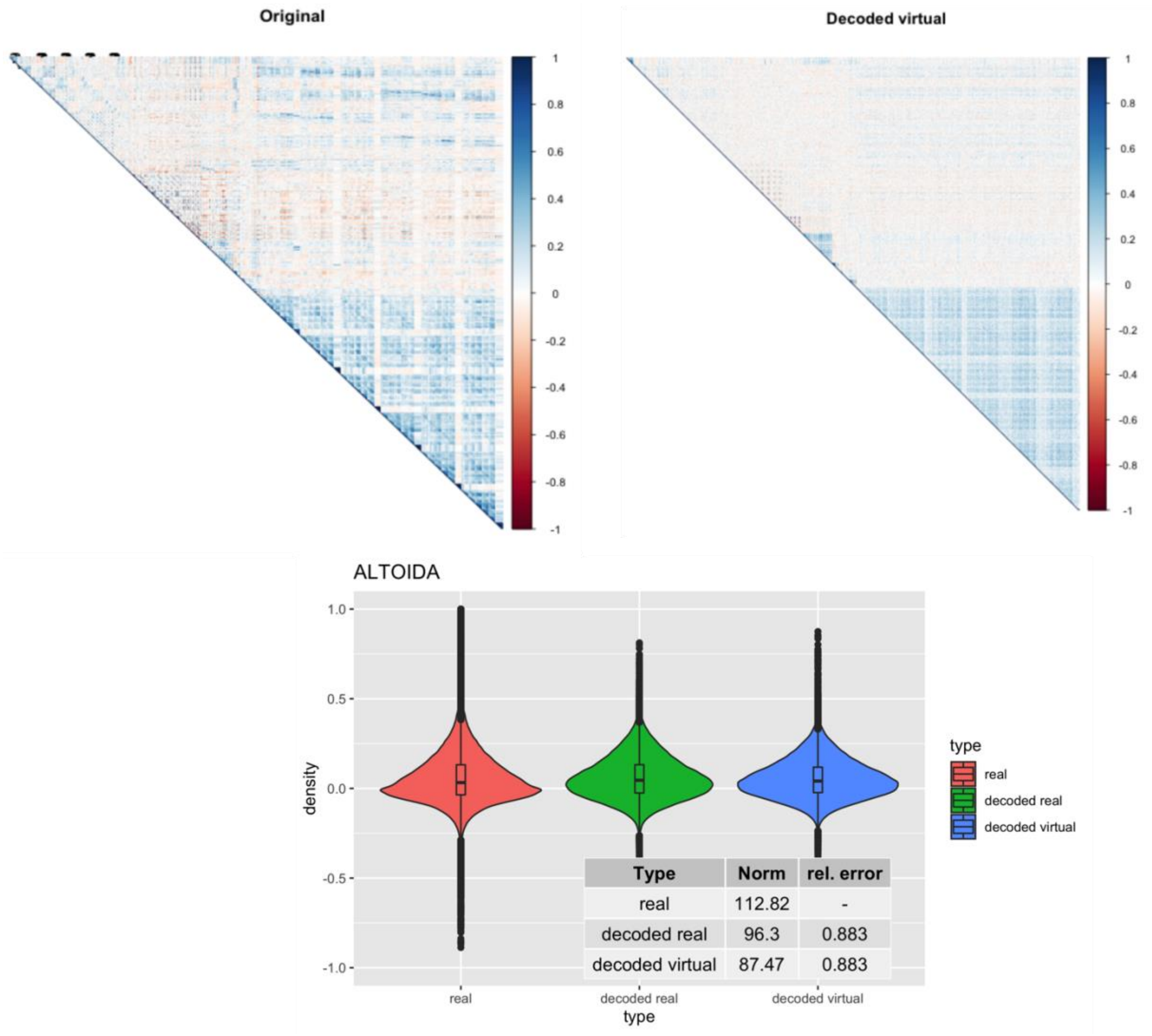

FIGURE S10. Top: Heatmaps reflecting the Pearson correlation matrix between variables in data and data generated via the VAMBN model trained on Altoida data.

Bottom: Distribution of Pearson correlation coefficients in real data (red), real patients (green) and decoded virtual / simulated subjects (blue). Tables show the Frobenius norm of the correlation matrices as well as the relative error, which consist of the norm of the matrix that is the difference between the real and decoded correlation matrix divided by the norm of the original correlation matrix.

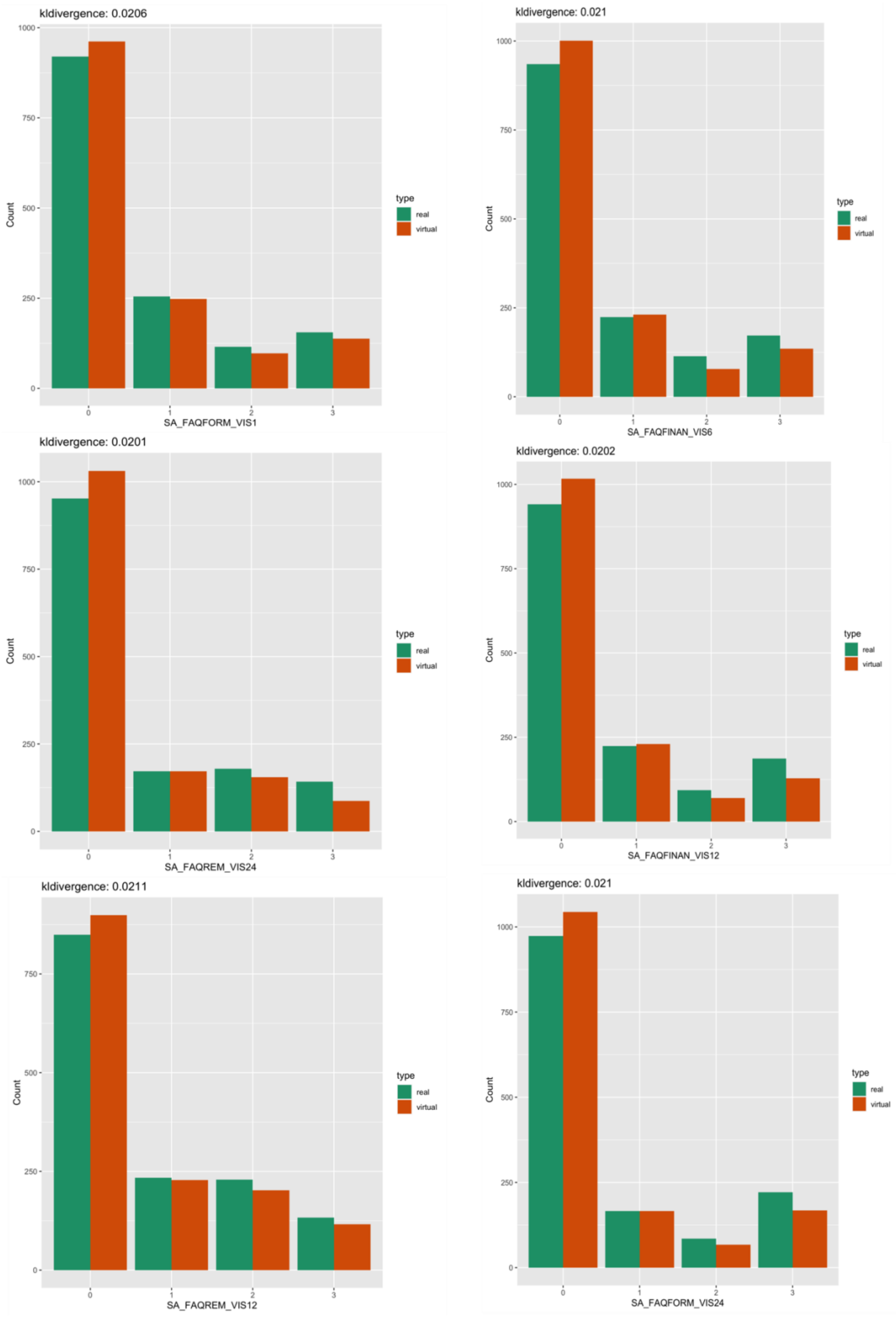

FIGURE S11. Examples of real and simulated patients for ADNI Data. The Figure compares the marginal distributions of FAQ variables for real patients (green) and virtual / generated patients (red). KL-divergence between the real and virtual patients is mentioned on the top of each plot.

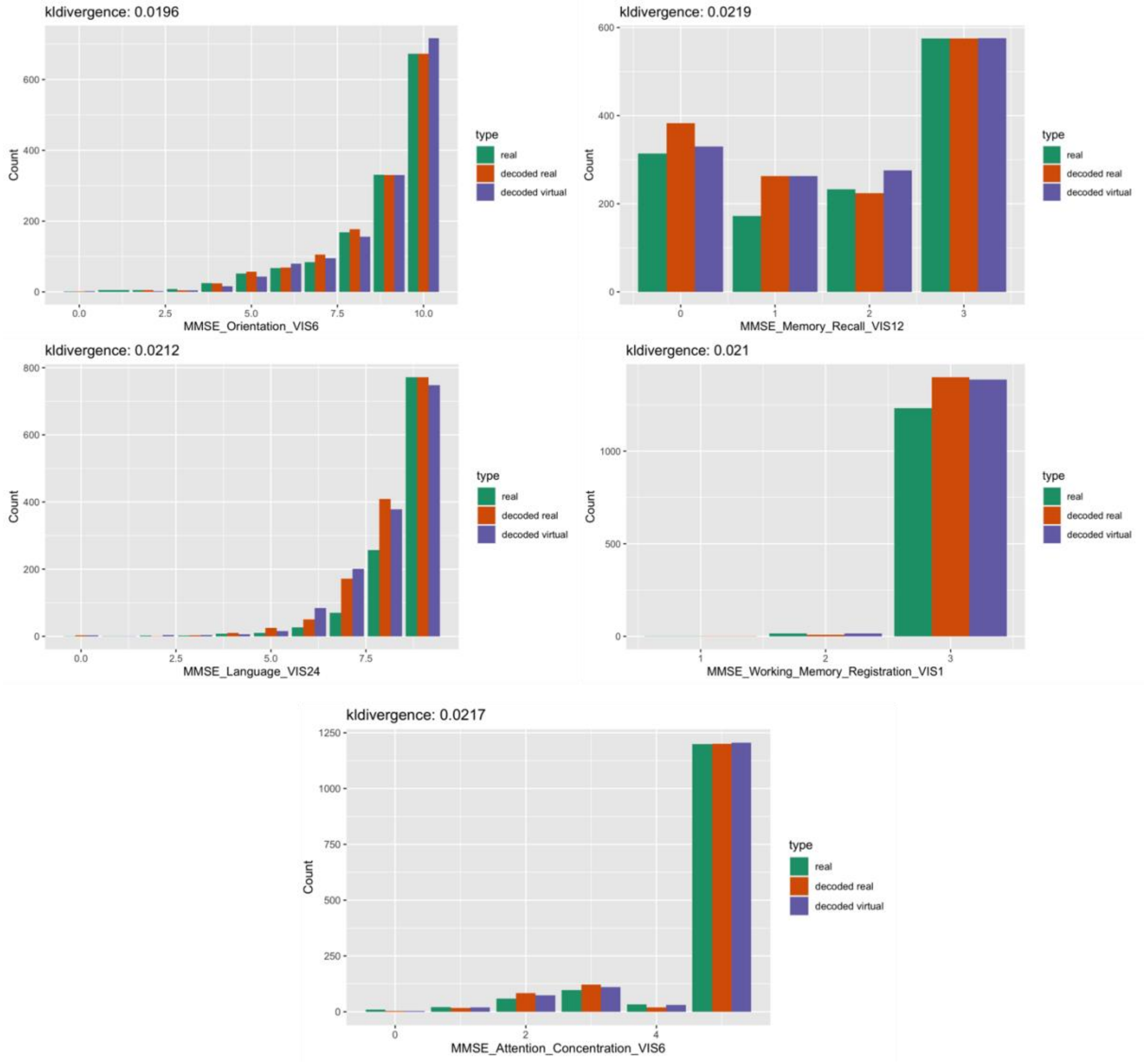

FIGURE S12. Examples of real and simulated patients for ADNI Data. The Figure compares the distributions of variables related to MMSE scores for real patients (green), virtual / generated patients (purple) and real patients decoded via the HI-VAE model (red). KL-divergence between the decoded real and decoded virtual patients is mentioned on the top of each plot.

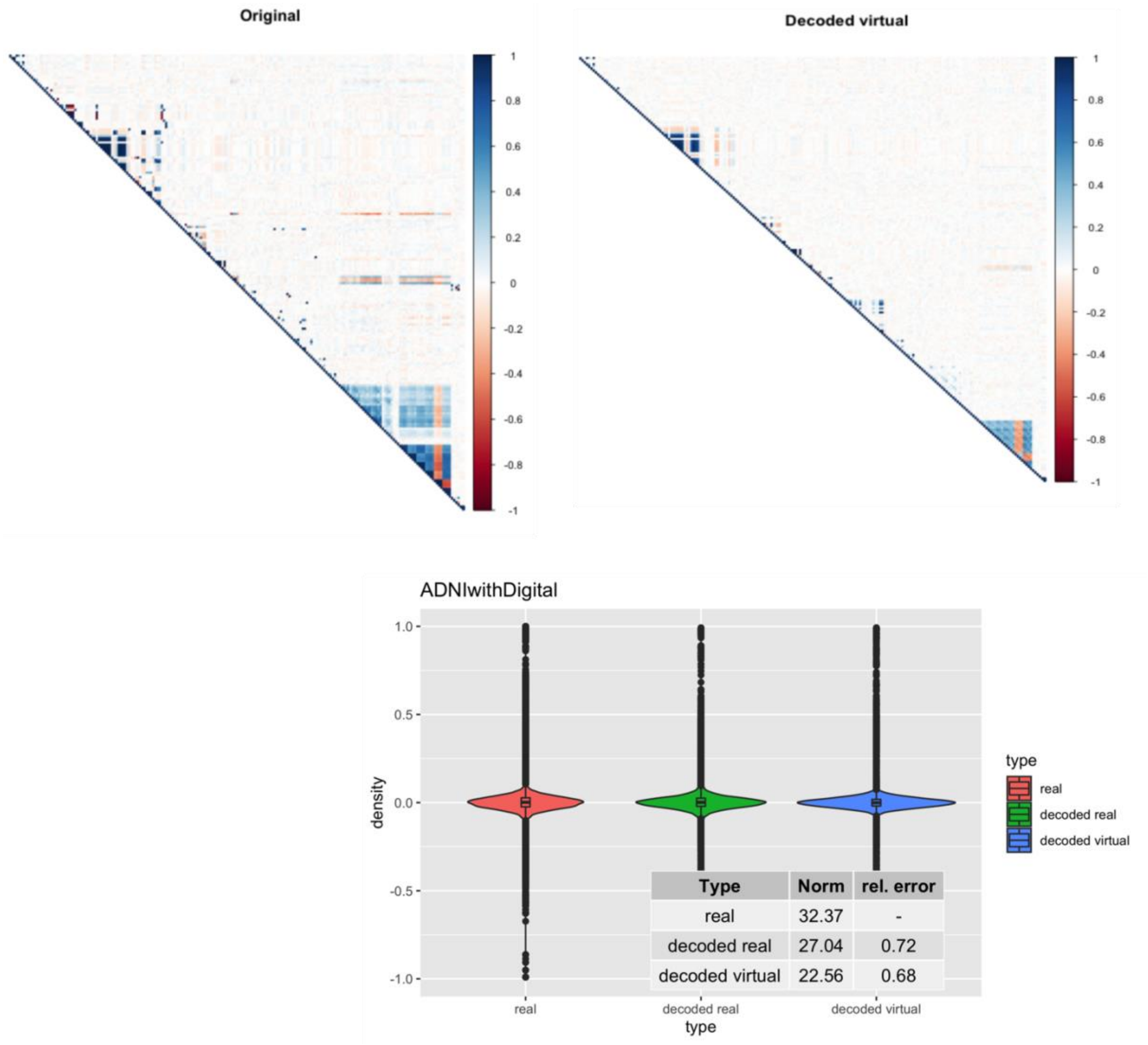

FIGURE S13. Top: Heatmaps reflecting the pearson Pearson correlation matrix between variables in real patients' data and decoded virtual patients' data generated via the VAMBN model trained on in ADNI data. Bottom: Distribution of pearson Pearson correlation coefficients between variables in real patients (red), decoded real patients (green) and virtual / simulated patients subjects (blue). Tables show the Frobenius norm of the correlation matrices as well as the relative error, which consist of the norm of the matrix that is the difference between the real and decoded correlation matrix dividedby the norm of the original correlation matrix.

#### 10. Sparse group lasso classifier

A sparse group lasso classifier depicts a linear combination between lasso and group lasso and provides sparse solution between both within and between groups[11]. It fits a regularized general linear model via penalized maximum likelihood. This method is used in cases where usually the number of features exceed the number of observations, and each feature group has varied number of variables[12]. Let $D = \{(x_i, y_i) | x_i \in \mathbb{R}^p, y_i \in \{0,1\}\}$ be the training data and let $m$ be the number of known groups in the data (e.g., DMs, MMSE scores). The number of features in the $l$th group is denoted by $p_l$. The sparse group lasso classifier solves the following optimization problem:

$$\beta^* = \arg\min_\beta \frac{1}{2n} \sum_{i=1}^{n} (y - g^{-1}(x_i^T \beta))^2 + (1-\alpha)\lambda \sum_{l=1}^{m} \sqrt{p_l} \left\| \beta^{(l)} \right\|_2 + \alpha\lambda \|\beta\|_1$$

where $g$ denotes the link function and $\beta^{(l)}$ the coefficients for features in the $l$th group. Regularization parameter $\lambda$ controls the overall amount of sparsity of the solution $\beta^*$, whereas $\alpha$ defines a trade-off between the lasso like $\ell_1$ penalty and the group lasso penalty, which favors a sparse selection of entire feature groups.
A grid search was performed to select the best set of hyperparameters $\alpha$ and $\lambda$.
This was done using a 10-fold cross-validation, which was nested inside the 5-fold cross-validation loop. The values considered for $\alpha$ was from 0.1 to 1 at an interval of 0.1, and 20 values for $\lambda$. The variables having no group structure were trained using simple lasso where $\alpha$ = 1 and best $\lambda$ was chosen from 20 $\lambda$ values. After selecting the best parameters, the performance of the model was evaluated using 5-fold cross validations repeated 10 times. In practice we used the R-package SGL for training, which automatically selected a range of values for $\lambda$.

##### 10.1. Sensitivity of DMs compared to clinical outcomes

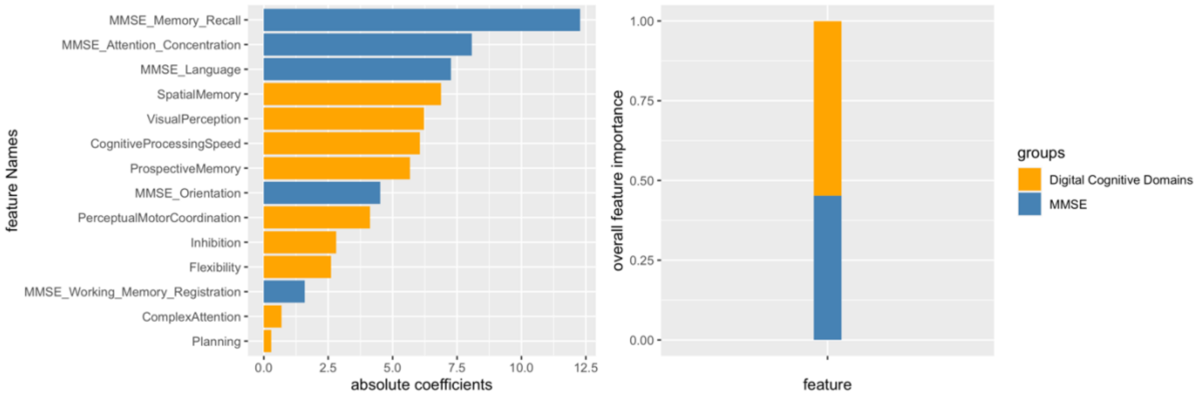

FIGURE S14. Feature importances observed in Altoida data (measured by the absolute value of coefficients) using a machine learning classifier discriminating between CN and MCI subjects. Left: feature importance when using MMSE plus digital cognitive domains. Right: overall feature importance of MMSE versus digital cognitive domains, measured by the sum of absolute coefficient values. The total sum of all absolute coefficient values is normalized to 1.

**TABLE S13. Feature Importances of classifier trained on MMSE and Digital Tasks**

| **Features** | **coef** | **abscoef** | **Group they belong to** |
|---|---|---|---|
| MMSE_Memory_Recall | -6.9048611612 | 6.9048611612 | MMSE |
| MMSE_Attention_Concentration | -5.2231597454 | 5.2231597454 | MMSE |
| Feature 1037 | -3.8849624365 | 3.8849624365 | ARObjectPlacement |
| MMSE_Orientation | -3.8057363479 | 3.8057363479 | MMSE |
| Feature 152 | -2.1365315989 | 2.1365315989 | MotorTappingFeatures |
| Feature 953 | -1.5298048803 | 1.5298048803 | ARObjectFinding |
| Feature 552 | -1.2494344228 | 1.2494344228 | ARObjectPlacement |
| Feature 271 | -1.1705960106 | 1.1705960106 | MotorTestDurations |
| Feature 555 | -1.1393259744 | 1.1393259744 | ARObjectPlacement |
| Feature 160 | -1.0917716097 | 1.0917716097 | MotorTappingFeatures |

| Features | coef | abscoef | Group they belong to |
|---|---|---|---|
| Feature 316 | -1.0634325025 | 1.0634325025 | MotorTestDurations |
| Feature 129 | -1.0608316922 | 1.0608316922 | ARObjectPlacement |
| Feature 127 | -0.9383948640 | 0.9383948640 | ARObjectFinding |
| Feature 297 | -0.8793787579 | 0.8793787579 | MotorTestDurations |
| Feature 550 | -0.8758272626 | 0.8758272626 | ARObjectPlacement |
| Feature 286 | -0.8522323588 | 0.8522323588 | MotorTestDurations |
| Feature 98 | -0.7988185690 | 0.7988185690 | ARGlobalTelemetryVariance |
| Feature 543 | -0.7106567297 | 0.7106567297 | ARObjectPlacement |
| Feature 277 | -0.6530482428 | 0.6530482428 | MotorTestDurations |
| Feature 310 | -0.6101853887 | 0.6101853887 | MotorTestDurations |
| Feature 930 | -0.5901836301 | 0.5901836301 | MotorTappingFeatures |
| Feature 922 | -0.5112133402 | 0.5112133402 | MotorTappingFeatures |
| Feature 553 | -0.4806854907 | 0.4806854907 | ARObjectPlacement |
| Feature 262 | -0.4295429135 | 0.4295429135 | MotorTestDurations |
| Feature 547 | -0.3780549645 | 0.3780549645 | ARObjectPlacement |
| Feature 929 | -0.1633435033 | 0.1633435033 | MotorTappingFeatures |
| Feature 313 | -0.1483455232 | 0.1483455232 | MotorTestDurations |
| Feature 591 | -0.0537978226 | 0.0537978226 | ARObjectPlacementFFT |
| Feature 628 | -0.0481819730 | 0.0481819730 | ARObjectPlacementFFT |
| Feature 655 | -0.0480896302 | 0.0480896302 | ARObjectPlacementFFT |
| Feature 830 | -0.0480151135 | 0.0480151135 | ARObjectPlacementFFT |
| Feature 581 | -0.0452055894 | 0.0452055894 | ARObjectPlacementFFT |
| Feature 802 | -0.0430511176 | 0.0430511176 | ARObjectPlacementFFT |
| Feature 598 | -0.0429720890 | 0.0429720890 | ARObjectPlacementFFT |
| Feature 769 | -0.0420179029 | 0.0420179029 | ARObjectPlacementFFT |
| Feature 654 | -0.0407232721 | 0.0407232721 | ARObjectPlacementFFT |
| Feature 829 | -0.0398399396 | 0.0398399396 | ARObjectPlacementFFT |
| Feature 890 | -0.0383355038 | 0.0383355038 | ARObjectPlacementFFT |

| Features | coef | abscoef | Group they belong to |
|---|---|---|---|
| Feature 782 | -0.0382514378 | 0.0382514378 | ARObjectPlacementFFT |
| Feature 775 | -0.0376142318 | 0.0376142318 | ARObjectPlacementFFT |
| Feature 562 | -0.0354050354 | 0.0354050354 | ARObjectPlacementFFT |
| Feature 592 | -0.0352397729 | 0.0352397729 | ARObjectPlacementFFT |
| Feature 661 | -0.0352116186 | 0.0352116186 | ARObjectPlacementFFT |
| Feature 599 | -0.0351937757 | 0.0351937757 | ARObjectPlacementFFT |
| Feature 590 | -0.0351608287 | 0.0351608287 | ARObjectPlacementFFT |
| Feature 893 | -0.0350350956 | 0.0350350956 | ARObjectPlacementFFT |
| Feature 571 | -0.0321512280 | 0.0321512280 | ARObjectPlacementFFT |
| Feature 579 | -0.0318804979 | 0.0318804979 | ARObjectPlacementFFT |
| Feature 650 | -0.0313653703 | 0.0313653703 | ARObjectPlacementFFT |
| Feature 828 | -0.0311762626 | 0.0311762626 | ARObjectPlacementFFT |
| Feature 894 | -0.0305373609 | 0.0305373609 | ARObjectPlacementFFT |
| Feature 580 | -0.0296295165 | 0.0296295165 | ARObjectPlacementFFT |
| Feature 858 | -0.0286589225 | 0.0286589225 | ARObjectPlacementFFT |
| Feature 910 | -0.0285759449 | 0.0285759449 | ARObjectPlacementFFT |
| Feature 738 | -0.0284599951 | 0.0284599951 | ARObjectPlacementFFT |
| Feature 917 | -0.0284131038 | 0.0284131038 | ARObjectPlacementFFT |
| Feature 914 | -0.0271049551 | 0.0271049551 | ARObjectPlacementFFT |
| Feature 784 | -0.0270741851 | 0.0270741851 | ARObjectPlacementFFT |
| Feature 594 | -0.0265930825 | 0.0265930825 | ARObjectPlacementFFT |
| Feature 711 | -0.0259945370 | 0.0259945370 | ARObjectPlacementFFT |
| Feature 724 | -0.0255489909 | 0.0255489909 | ARObjectPlacementFFT |
| Feature 605 | -0.0254415974 | 0.0254415974 | ARObjectPlacementFFT |
| Feature 609 | -0.0254369701 | 0.0254369701 | ARObjectPlacementFFT |
| Feature 700 | -0.0253632350 | 0.0253632350 | ARObjectPlacementFFT |
| Feature 913 | -0.0252823528 | 0.0252823528 | ARObjectPlacementFFT |
| Feature 678 | -0.0250451628 | 0.0250451628 | ARObjectPlacementFFT |

| Features | coef | abscoef | Group they belong to |
|---|---|---|---|
| Feature 589 | -0.0243233995 | 0.0243233995 | ARObjectPlacementFFT |
| Feature 607 | -0.0235596032 | 0.0235596032 | ARObjectPlacementFFT |
| Feature 652 | -0.0231908988 | 0.0231908988 | ARObjectPlacementFFT |
| Feature 831 | -0.0227642826 | 0.0227642826 | ARObjectPlacementFFT |
| Feature 785 | -0.0221150050 | 0.0221150050 | ARObjectPlacementFFT |
| Feature 761 | -0.0220418675 | 0.0220418675 | ARObjectPlacementFFT |
| Feature 911 | -0.0220184438 | 0.0220184438 | ARObjectPlacementFFT |
| Feature 710 | -0.0218650339 | 0.0218650339 | ARObjectPlacementFFT |
| Feature 569 | -0.0218308983 | 0.0218308983 | ARObjectPlacementFFT |
| Feature 781 | -0.0209373460 | 0.0209373460 | ARObjectPlacementFFT |
| Feature 611 | -0.0208605979 | 0.0208605979 | ARObjectPlacementFFT |
| Feature 586 | -0.0206759723 | 0.0206759723 | ARObjectPlacementFFT |
| Feature 606 | -0.0203051032 | 0.0203051032 | ARObjectPlacementFFT |
| Feature 727 | -0.0200738528 | 0.0200738528 | ARObjectPlacementFFT |
| Feature 574 | -0.0197875811 | 0.0197875811 | ARObjectPlacementFFT |
| Feature 640 | -0.0187237830 | 0.0187237830 | ARObjectPlacementFFT |
| Feature 715 | -0.0186184563 | 0.0186184563 | ARObjectPlacementFFT |
| Feature 725 | -0.0174915438 | 0.0174915438 | ARObjectPlacementFFT |
| Feature 714 | -0.0166338940 | 0.0166338940 | ARObjectPlacementFFT |
| Feature 776 | -0.0166243569 | 0.0166243569 | ARObjectPlacementFFT |
| Feature 912 | -0.0166155075 | 0.0166155075 | ARObjectPlacementFFT |
| Feature 563 | -0.0164072313 | 0.0164072313 | ARObjectPlacementFFT |
| Feature 596 | -0.0163643362 | 0.0163643362 | ARObjectPlacementFFT |
| Feature 578 | -0.0162497237 | 0.0162497237 | ARObjectPlacementFFT |
| Feature 608 | -0.0161937892 | 0.0161937892 | ARObjectPlacementFFT |
| Feature 859 | -0.0159193561 | 0.0159193561 | ARObjectPlacementFFT |
| Feature 783 | -0.0154647638 | 0.0154647638 | ARObjectPlacementFFT |
| Feature 916 | -0.0152167915 | 0.0152167915 | ARObjectPlacementFFT |

| Features | coef | abscoef | Group they belong to |
|---|---|---|---|
| Feature 891 | -0.0151750551 | 0.0151750551 | ARObjectPlacementFFT |
| Feature 720 | -0.0151197165 | 0.0151197165 | ARObjectPlacementFFT |
| Feature 618 | -0.0149766466 | 0.0149766466 | ARObjectPlacementFFT |
| Feature 595 | -0.0147348993 | 0.0147348993 | ARObjectPlacementFFT |
| Feature 869 | -0.0147062679 | 0.0147062679 | ARObjectPlacementFFT |
| Feature 651 | -0.0142790657 | 0.0142790657 | ARObjectPlacementFFT |
| Feature 593 | -0.0142154924 | 0.0142154924 | ARObjectPlacementFFT |
| Feature 915 | -0.0141376845 | 0.0141376845 | ARObjectPlacementFFT |
| Feature 614 | -0.0139681272 | 0.0139681272 | ARObjectPlacementFFT |
| Feature 617 | -0.0139583111 | 0.0139583111 | ARObjectPlacementFFT |
| Feature 664 | -0.0139241247 | 0.0139241247 | ARObjectPlacementFFT |
| Feature 838 | -0.0133645027 | 0.0133645027 | ARObjectPlacementFFT |
| Feature 656 | -0.0132535866 | 0.0132535866 | ARObjectPlacementFFT |
| Feature 812 | -0.0131829465 | 0.0131829465 | ARObjectPlacementFFT |
| Feature 903 | -0.0130345092 | 0.0130345092 | ARObjectPlacementFFT |
| Feature 888 | -0.0129726731 | 0.0129726731 | ARObjectPlacementFFT |
| Feature 610 | -0.0123856080 | 0.0123856080 | ARObjectPlacementFFT |
| Feature 842 | -0.0121619010 | 0.0121619010 | ARObjectPlacementFFT |
| Feature 758 | -0.0108101736 | 0.0108101736 | ARObjectPlacementFFT |
| Feature 653 | -0.0099864470 | 0.0099864470 | ARObjectPlacementFFT |
| Feature 663 | -0.0097990729 | 0.0097990729 | ARObjectPlacementFFT |
| Feature 548 | -0.0092283166 | 0.0092283166 | ARObjectPlacement |
| Feature 625 | -0.0091881468 | 0.0091881468 | ARObjectPlacementFFT |
| Feature 689 | -0.0088471857 | 0.0088471857 | ARObjectPlacementFFT |
| Feature 774 | -0.0087969288 | 0.0087969288 | ARObjectPlacementFFT |
| Feature 588 | -0.0087344845 | 0.0087344845 | ARObjectPlacementFFT |
| Feature 754 | -0.0082567409 | 0.0082567409 | ARObjectPlacementFFT |
| Feature 702 | -0.0082383752 | 0.0082383752 | ARObjectPlacementFFT |

| Features | coef | abscoef | Group they belong to |
|---|---|---|---|
| Feature 603 | -0.0082290682 | 0.0082290682 | ARObjectPlacementFFT |
| Feature 832 | -0.0081682944 | 0.0081682944 | ARObjectPlacementFFT |
| Feature 872 | -0.0074034436 | 0.0074034436 | ARObjectPlacementFFT |
| Feature 602 | -0.0066559038 | 0.0066559038 | ARObjectPlacementFFT |
| Feature 909 | -0.0065067260 | 0.0065067260 | ARObjectPlacementFFT |
| Feature 773 | -0.0063594579 | 0.0063594579 | ARObjectPlacementFFT |
| Feature 683 | -0.0062824507 | 0.0062824507 | ARObjectPlacementFFT |
| Feature 662 | -0.0060786495 | 0.0060786495 | ARObjectPlacementFFT |
| Feature 823 | -0.0057044472 | 0.0057044472 | ARObjectPlacementFFT |
| Feature 712 | -0.0056766156 | 0.0056766156 | ARObjectPlacementFFT |
| Feature 760 | -0.0054441671 | 0.0054441671 | ARObjectPlacementFFT |
| Feature 660 | -0.0048696732 | 0.0048696732 | ARObjectPlacementFFT |
| Feature 892 | -0.0045558408 | 0.0045558408 | ARObjectPlacementFFT |
| Feature 849 | -0.0036718923 | 0.0036718923 | ARObjectPlacementFFT |
| Feature 854 | -0.0035120333 | 0.0035120333 | ARObjectPlacementFFT |
| Feature 868 | -0.0033360821 | 0.0033360821 | ARObjectPlacementFFT |
| Feature 757 | -0.0032630884 | 0.0032630884 | ARObjectPlacementFFT |
| Feature 853 | -0.0032531313 | 0.0032531313 | ARObjectPlacementFFT |
| Feature 719 | -0.0032391173 | 0.0032391173 | ARObjectPlacementFFT |
| Feature 850 | -0.0031009246 | 0.0031009246 | ARObjectPlacementFFT |
| Feature 613 | -0.0030885988 | 0.0030885988 | ARObjectPlacementFFT |
| Feature 780 | -0.0030709555 | 0.0030709555 | ARObjectPlacementFFT |
| Feature 851 | -0.0028858108 | 0.0028858108 | ARObjectPlacementFFT |
| Feature 848 | -0.0028712393 | 0.0028712393 | ARObjectPlacementFFT |
| Feature 855 | -0.0027249662 | 0.0027249662 | ARObjectPlacementFFT |
| Feature 657 | -0.0026265114 | 0.0026265114 | ARObjectPlacementFFT |
| Feature 889 | -0.0023347196 | 0.0023347196 | ARObjectPlacementFFT |
| Feature 561 | -0.0017700738 | 0.0017700738 | ARObjectPlacementFFT |

| Features | coef | abscoef | Group they belong to |
|---|---|---|---|
| Feature 722 | -0.0014810635 | 0.0014810635 | ARObjectPlacementFFT |
| Feature 852 | -0.0012712528 | 0.0012712528 | ARObjectPlacementFFT |
| Feature 857 | -0.0006237352 | 0.0006237352 | ARObjectPlacementFFT |

**10.2 Sensitivity of FAQ compared to MMSE**

For ADNI, we focused on the baseline data of subjects that maintained their diagnostic state from study baseline till month 24. This resulted into 360 CN, 403 MCI and 147 demented subjects. Since the prediction of digital measures for ADNI patients depends on the diagnostic status, we only considered the following combination of cognition scores:

1. MMSE subitem scores and FAQ subitem scores
2. MMSE subitem scores
3. FAQ subitem scores

**10.3 Comparison of diagnostic value of FAQ and MMSE in ADNI**

Our results on ADNI demonstrated a significant increase in the prediction performance on held out test data when using FAQ in addition to MMSE subitem scores ($p<0.0001$, Wilcoxon signed rank test), compared to only using MMSE scores. Similarly, when using FAQ scores alone the prediction performance was significantly better compared to only using MMSE scores ($p=0.04$, Wilcoxon signed rank test).

We also explored the absolute values of the coefficients in the classifier using MMSE plus FAQ scores. The most relevant MMSE score was the domain of “memory and recall” followed by FAQ subitem score related to “remembering appointments, occasions etc.” and “assembling tax records, business affairs”. This analysis demonstrated that FAQ subitem scores contributed 78.7% of the overall feature importance, whereas MMSE subitem scores contributed 21.3% (Figure S15).

A

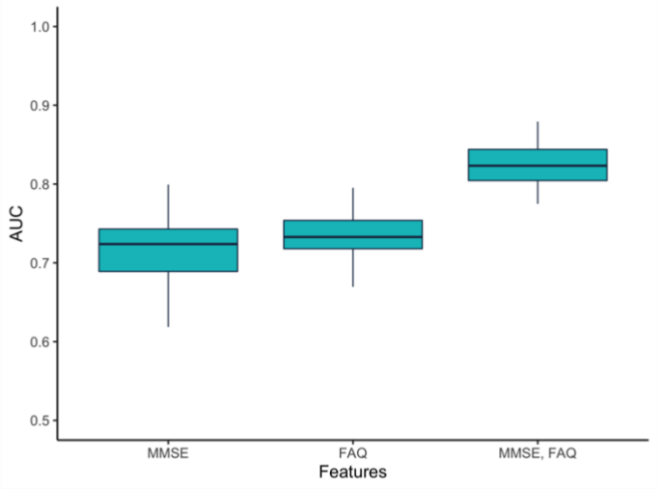

B

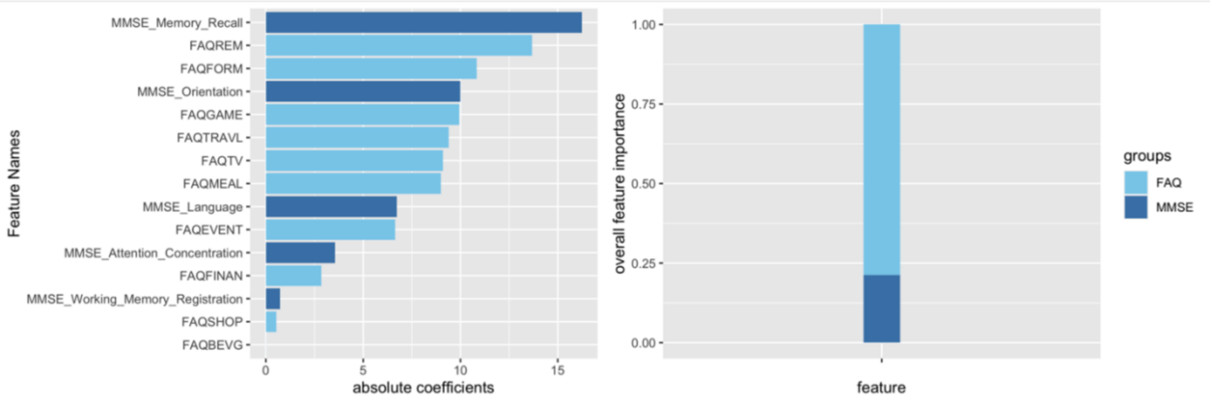

FIGURE S15. A: Performance of a sparse group lasso trained on different modalities of data to classify subjects into CN and MCI using ADNI data, measured via the area under ROC curve (AUC-ROC). The boxplots show the distribution of the AUC-ROC on held out test data, measured via 10 times repeated 5-fold repeated cross-validation.
B: Feature importance when using MMSE plus individual FAQ observed in ADNI data (measured by the absolute value of coefficients) using a machine learning classifier discriminating between CN and MCI patients, overall feature importance of MMSE versus FAQ, measured by the sum of absolute coefficient values. The total sum of all absolute coefficient values is normalized to 1.

**10.4 External validation of classifier trained on ADNI MMSE features**

We externally validated the prediction performance of the sparse group lasso classifier trained on ADNI MMSE data by testing it on Altoida data. A slight drop in the prediction performance to an AUC of 82% was observed (Figure S16) compared to the 10-fold cross-validated AUC of ~86% using Altoida data

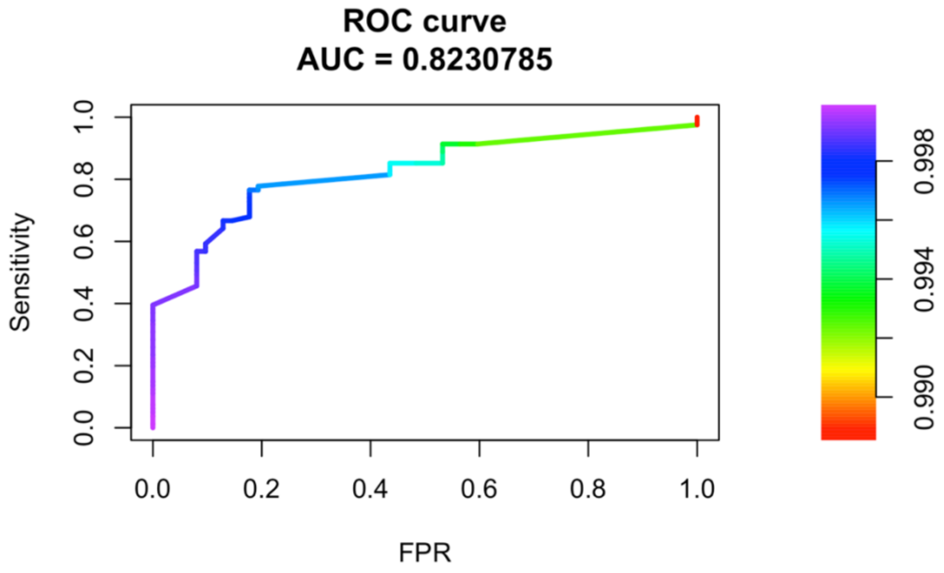

FIGURE S16. Performance of classifier trained with MMSE subitem scores using ADNI data and tested on Altoida data.
